# Time-aware machine learning enables early intrapartum identification of fetuses at risk of hypoxic-ischemic encephalopathy from cardiotocography

**DOI:** 10.64898/2026.09.28.26364144

**Authors:** Johann Vargas-Calixto, Michael W. Kuzniewicz, Marie-Coralie Cornet, Yvonne W. Wu, Aditi Lahiri, Lawrence Gerstley, John Parker, Philip A. Warrick, Robert E. Kearney

## Abstract

Cardiotocography (CTG) is the standard of care for intrapartum fetal monitoring, yet its poor specificity drives unnecessary intervention without reducing hypoxic-ischemic encephalopathy (HIE). Most artificial intelligence (AI) models are trained exclusively on the final hour before delivery, a window only identifiable retrospectively. We hypothesized that this framework prevents models from learning the early evolving signs of HIE. We analyzed 174, 186 deliveries, with gestational age GA ≥ 35 weeks, from Kaiser Permanente Northern California. Forty CTG features were extracted from 20-minute epochs to train Random Forest classifiers targeting severe acidosis (***N*** = 2, 636) and clinically validated HIE (***N*** = 304). The training window was progressively extended from the final hour to the full duration of labor, and time from labor onset (TLO) was added as a feature. Models were evaluated continuously throughout labor at a fixed 15% false positive rate. Our results showed that the early identification of fetuses at risk of HIE improved as the training window was extended, plateauing at 18 hours. Adding TLO gave the highest early-warning performance: 40.3% of HIE cases identified at least 3 hours and 27.3% at least 6 hours before delivery, absolute gains of 8.4% and 10.7% over the last-hour classifier. Finally, 55.7% of HIE cases were identified at least 40 minutes before delivery. In conclusion, classifiers trained only on the last hour may identify HIE patterns that occur near delivery; time-aware models trained across labor map the temporal evolution of these patterns and provide actionable early warnings. Thus, AI systems for intrapartum monitoring must account for the temporal evolution of labor to shift from end-of-labor diagnosis to early-warning decision support.

**Author summary:** During labor, fetal monitoring relies on cardiotocography (CTG) to assess fetal well-being and their risk of developing hypoxic-ischemic encephalopathy (HIE). However, current clinical guidelines suffer from poor specificity, driving unnecessary Caesarean deliveries without decreasing HIE rates. Moreover, existing artificial intelligence (AI) systems are trained exclusively on the final hour of labor, which is only available retrospectively and assume signal stationarity. The fetal heart rate (FHR) evolves continuously through labor, rendering the assumption of stationarity reductive. This biased assumption also limits the early-warning capabilities of such AI systems. Here, we analyzed 174,186 deliveries to train random-forest (RF) models that target severe acidosis and HIE. We found that expanding the training window beyond the final hour and indexing the time of labor onset into the classification features brought significant advantage. This joint approach resulted in over 10.7% more HIE cases being detected at six hours before delivery when compared to the conventional last-hour approach. Such achievement carries important clinical implications. For preventive interventions to be appropriate and efficient, clinicians must be able to stratify risk as early as possible. Thus, the goal of the intrapartum decision support systems should not be limited to accurate detections at the time of delivery but also to optimize for the lead time of the detections. Our approach demonstrated that to significantly improve the early identification of fetuses at risk of HIE, novel AI systems should be exposed to longer labor training windows and be able to track the time from labor onset to provide temporal context to their decisions.

## 1 Introduction

Intrapartum hypoxic-ischemic encephalopathy (HIE) is an adverse perinatal outcome with catastrophic consequences; severe HIE contributes significantly to neonatal mortality and lifelong neurodevelopmental morbidity, including cerebral palsy [1]. During labor, the fetus is subjected to intermittent hypoxic stress. Uterine contractions temporarily reduce blood flow to the placenta and umbilical cord, impairing gas exchange, so the fetus must rely on its oxygen reserves and compensatory mechanisms to maintain cerebral perfusion [2,3]. Excessive uterine contractions, placental insufficiency, or predisposed fetal vulnerability may overwhelm these mechanisms, leading to progressive physiological deterioration and worsening metabolic acidosis. Ultimately, without recovery or timely fetal rescue, brain-sparing mechanisms fail, and the fetus sustains cerebral injury defined as HIE [4].

Since its introduction over 50 years ago, cardiotocography (CTG) has been the primary tool for intrapartum fetal monitoring. CTG comprises the visual assessment of fetal heart rate (FHR) and uterine activity (UA) tracings to identify fetuses at risk of intrapartum compromise. However, visual CTG interpretation carries high intra- and inter-observer variability [5]. Moreover, clinical guidelines for CTG interpretation lack specificity, with over 80% of tracings exhibiting indeterminate patterns during labor [6]. This limitation has led to paradoxical intervention outcomes: continuous CTG monitoring increases the rate of Caesarean deliveries without a corresponding decrease in the incidence of cerebral palsy or perinatal mortality [7], while the incidence of HIE remains stable [8].

In response to these limitations, artificial intelligence (AI) research has pursued objective and highly sensitive algorithms to detect fetuses at increased risk of HIE. Although numerous AI models report strong retrospective performance, none have translated to routine intrapartum care. A recent comprehensive consensus report highlighted this gap, calling for strategic initiatives to close it [9]. That gap has several contributing causes, and we address two of them. The first is the use of weak surrogate outcomes as labels; most models are trained against lenient biochemical thresholds instead of relying on confirmed clinical outcomes. In this study we target severe acidosis and the clinical diagnosis of HIE. The second, and the focus of this work, is that published models are anchored in time by design; these models are trained and evaluated on a specific period of labor relative to the time of delivery, often the last hour [10–13]. The time of delivery is not known during labor, so these models can only be scored retrospectively. We designed our models to not depend on the time of delivery during classification. Training data are selected retrospectively, when the time of delivery is known, but our resulting time-aware classifier only ever sees the current epoch and its time from labor onset. Thus, it can be run continuously throughout labor without needing to know when delivery will occur. We use time of delivery only as the axis along which the lead time of the predictions is measured, not as a model input. Table 1 provides a summary of recent high-impact AI approaches, highlighting these persisting constraints. The following subsections elaborate further on these gaps.

**Table 1.**
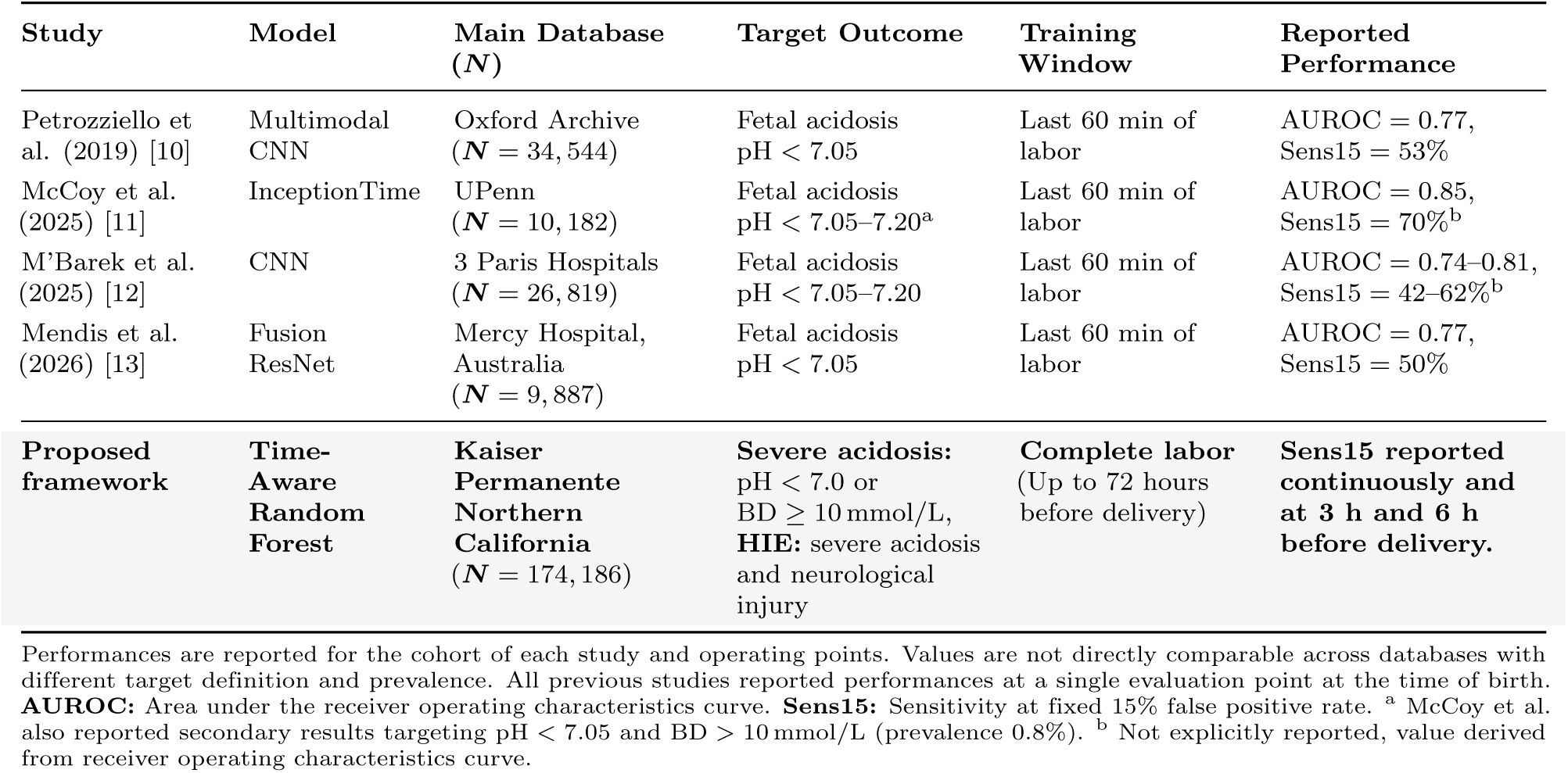
Comparison of key AI approaches against the proposed framework.

| Study | Model | Main Database<br>(*N*) | Target Outcome | Training Window | Reported Performance |
| --- | --- | --- | --- | --- | --- |
| Petrozziello et al. (2019) [10] | Multimodal CNN | Oxford Archive (*N* = 34,544) | Fetal acidosis<br>pH < 7.05 | Last 60 min of labor | AUROC = 0.77,<br>Sens15 = 53% |
| McCoy et al. (2025) [11] | InceptionTime | UPenn (*N* = 10,182) | Fetal acidosis<br>pH < 7.05–7.20<sup>a</sup> | Last 60 min of labor | AUROC = 0.85,<br>Sens15 = 70%<sup>b</sup> |
| M'Barek et al. (2025) [12] | CNN | 3 Paris Hospitals (*N* = 26,819) | Fetal acidosis<br>pH < 7.05–7.20 | Last 60 min of labor | AUROC = 0.74–0.81,<br>Sens15 = 42–62%<sup>b</sup> |
| Mendis et al. (2026) [13] | Fusion ResNet | Mercy Hospital, Australia (*N* = 9,887) | Fetal acidosis<br>pH < 7.05 | Last 60 min of labor | AUROC = 0.77,<br>Sens15 = 50% |
| **Proposed framework** | **Time-Aware Random Forest** | **Kaiser Permanente Northern California**<br>(*N* = 174,186) | **Severe acidosis:**<br>pH < 7.0 or<br>BD ≥ 10 mmol/L,<br>**HIE:** severe acidosis<br>and neurological injury | **Complete labor**<br>(Up to 72 hours before delivery) | **Sens15 reported continuously and at 3 h and 6 h before delivery.** |

Performances are reported for the cohort of each study and operating points. Values are not directly comparable across databases with different target definition and prevalence. All previous studies reported performances at a single evaluation point at the time of birth. **AUROC:** Area under the receiver operating characteristics curve. **Sens15:** Sensitivity at fixed 15% false positive rate. <sup>a</sup> McCoy et al. also reported secondary results targeting pH < 7.05 and BD > 10 mmol/L (prevalence 0.8%). <sup>b</sup> Not explicitly reported, value derived from receiver operating characteristics curve.

### 1.1 Biochemical acidosis as a surrogate for neurological injury

The vast majority of AI classification studies use umbilical cord arterial blood gases, most often pH alone, as the ground truth for fetal distress [10–14]. However, using biochemical acidosis alone, regardless of which blood-gas index is used, is an incomplete proxy for the more clinically relevant outcome: hypoxic-ischemic neurological injury.

During labor, intermittent hypoxia-ischemia depletes the oxygen reserves of the fetus, forcing a shift to anaerobic metabolism whose byproducts, e.g., lactic acid, deplete the fetal buffering system [15]. Although acidosis at birth measures the cumulative metabolic cost of labor, it does not confirm whether the same insult caused the hemodynamic failure and reduced cerebral perfusion that lead to HIE. Most infants that develop acidosis do not develop neurological injury [16].

If biochemical acidosis is the target outcome, the threshold that defines the target class becomes critical. Table 1 illustrates that most studies define intrapartum compromise for pH *<* 7.05, with some employing even more lenient cutoffs at pH *<* 7.20 [11,12]. However, the absolute risk of adverse neurologic outcomes is concentrated in the most severe cases of acidosis: 2.95% for pH *<* 7.0 falling to 0.36% as the target class widened to include all pH *<* 7.1 [16]. The 7.0 ≤ pH *<* 7.1 band is far larger than the pH *<* 7.0 band and carries considerably lower risk, so relaxing the threshold dilutes the risk of the target class. Also, mild acidosis above the 7.1 threshold is weakly associated with HIE or neonatal morbidity (risk below 0.16%) [16–18]. In most of these mild cases, the fetus successfully withstands labor. Ideally, AI classifiers should distinguish normal fetal stress from pathological fetal distress [19]. Otherwise, scaling these algorithms risks further increasing the already high Caesarean delivery rate. In this context, we propose targeting two outcomes, the clinical diagnosis of HIE and severe acidosis, in the development of AI classifiers.

### 1.2 The assumption of stationarity

The second constraint, and the more critical one, is the assumption of FHR stationarity that underlies temporal anchoring. As shown in Table 1, it is standard for AI models to be trained exclusively on the final hour of labor, immediately preceding birth [10–13]. Then, models are scored using the area under the receiver operating characteristics curve (AUROC) and sensitivity evaluated at this single time point. This end-of-labor framing hampers clinical applicability.

To effectively drive a decrease in the rates of neurological injury, AI systems must detect high-HIE-risk fetuses with sufficient lead time to allow for clinical intervention. The interval from the decision to intervene to an emergency Caesarean delivery – the standard emergency protocol for high risk cases – takes about 30 minutes, depending on hospital protocols and settings [20,21]. Thus, algorithms must provide early warnings in the order of hours and not minutes; identifying a high-risk case close to the time of birth offers no actionable preventive utility.

Moreover, FHR properties are not stationary. As uterine contractions intensify and cumulative labor stress increases, FHR properties evolve [22]. Fundamentally, AI classifiers identify pathological patterns in training data and seek similar patterns in new data during deployment. If the patterns in the signals are continuously evolving, it is unrealistic to expect a model trained exclusively on end-of-labor data to recognize early warnings of fetal distress. The continuous real-time deployment of such a temporally constrained model risks detecting only acute end-of-labor deterioration without identifying the early and persistent signs of intrapartum neurological injury.

### 1.3 Objectives and study rationale

The primary objective of this study is to systematically evaluate the impact of using temporal data on the early identification of fetuses at risk of intrapartum HIE. We previously showed that CTG features vary systematically through labor, and that their association with HIE is also time-varying [22]. Then, we showed that the time from labor onset (TLO), selected from a set of candidate time-surrogate variables, recovers that time-varying association prospectively, without requiring the time of delivery and could be used in future classification studies [23]. Here, we test whether these enhanced associations translate into improved early-warning classification performance under a realistic deployment evaluation that does not require knowing the time of delivery. We hypothesize that classifiers that explicitly incorporate the continuous evolution of labor will demonstrate superior early-warning capabilities compared to models that assume stationarity. To test this hypothesis, our experimental design is structured around three specific objectives:

1. To evaluate a standard end-of-labor classifier under idealized conditions versus continuous monitoring, quantifying its bias towards acute acidosis.
2. To determine how far the training window must extend beyond the last hour of labor to improve early identification of fetuses at risk of HIE.
3. To evaluate the additional predictive value of embedding the TLO directly into the classification feature space.

To carry out this evaluation, we deliberately held the model architecture fixed to a Random Forest (RF) throughout. The main limitation that we investigate is a property of the training data and not of the model architecture. Regardless of the number of trainable parameters, a model that never sees the hours of labor preceding delivery cannot learn patterns that occur there. For this reason, we keep the model architecture constant while we compare extending the training window and including TLO as a classification feature. RF suits our study for the following reasons. First, our design requires the training of 3,300 independent models: 10 × 5-fold cross-validated models, with 5 inner cross-validated classifiers per comparison. This number of trained models, across over 4.3 million CTG epochs, would be computationally prohibitive with models of larger capacity. Second, RF models require little hyperparameter tuning, facilitating comparisons across different training windows. Third, RF also handle severe class imbalance through per-tree weighting while avoiding synthetic resampling. Finally, RF facilitate CTG feature importance estimates, which leads to the time-resolved analysis in Section 3.4. While deep learning architectures might outperform our models, we are concerned with the direction and magnitude of the training window effect under a controlled comparison. Determining the architecture that best exploits the extended training window is a separate study.

## 2 Materials and methods

### 2.1 Ethics statement

This retrospective study analyzed clinical and CTG data collected between 2011 and 2019 from 15 Kaiser Permanente Northern California (KPNC) hospitals. The Research Ethics Boards of KPNC (approval numbers 1470283-1 and 1276201-29) and McGill University (approval numbers A04-M27-20A (20-04-025) and A04-M29-20B (20-04-056)) reviewed and approved the study. The study involved secondary research of data for which consent was not required. Data access was granted to all participating institutions starting September 25th 2020, and continuous data access has remained active for research purposed since then. For all records, personal health information was removed by KPNC personal before granting access for research purposes. Thus, the authors did not have access to any information that could enable the identification of the study participants.

### 2.2 Study population

There were 297,280 singleton live births at term and late preterm gestation (35–44 weeks) without major congenital or chromosomal anomalies at KPNC between 2011 and 2019. All available intrapartum CTG recordings were analyzed from as early as 72 hours before delivery to the time of delivery. Each record corresponds to one singleton delivery; we refer to this unit as an infant throughout.

Fig 1 shows the number of infants, and those selected for analysis. We reviewed all available CTG and electronic records to detect the onset of labor [24]. Cases in which labor onset could not be determined were excluded, along with the portions of the CTG signals recorded before labor onset.

**Fig 1.**
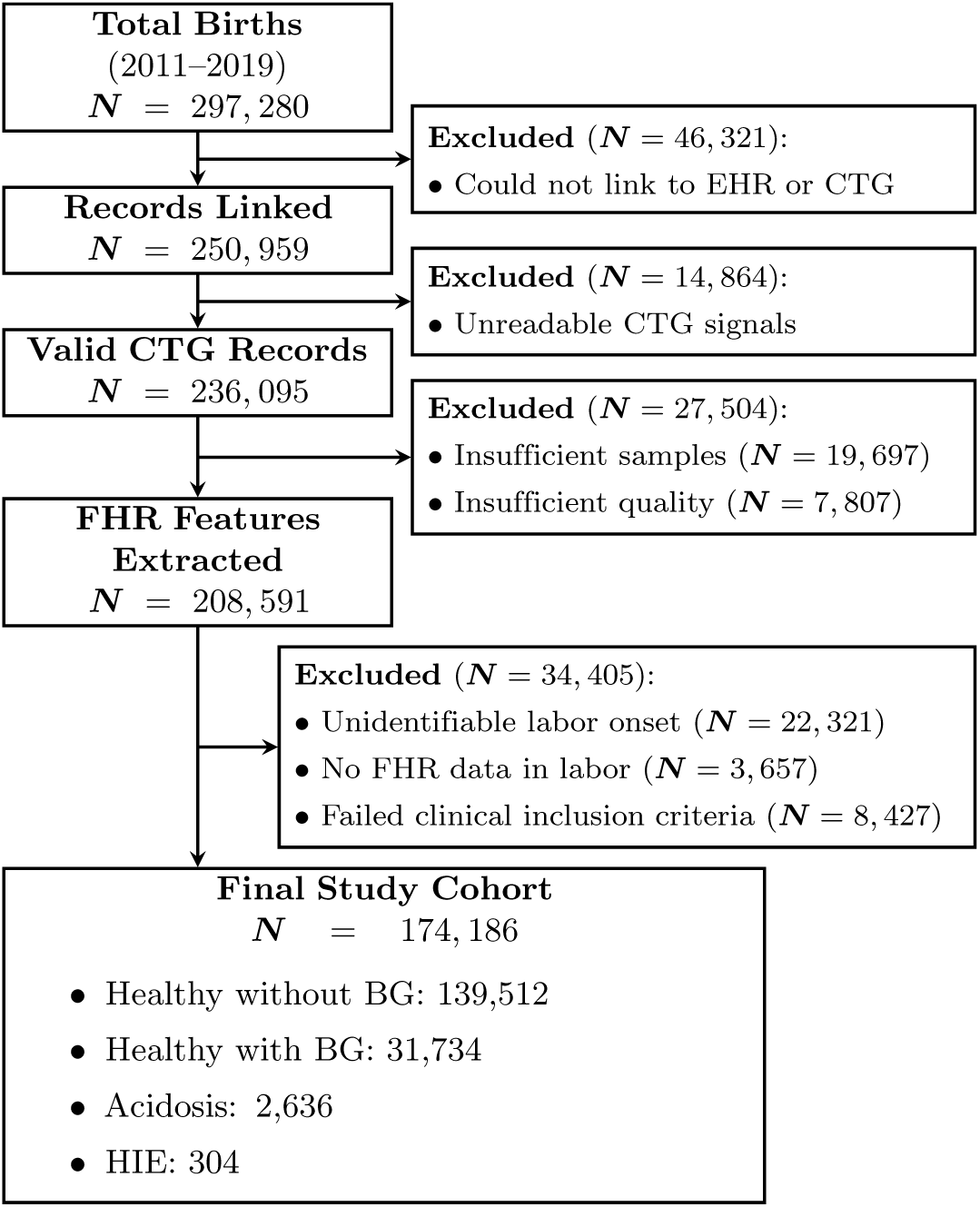
**Inclusion and exclusion criteria of the study cohort.**

#### 2.2.1 Labor outcomes definition

The outcomes in our cohort were defined based on the availability and the results of blood gas (BG) tests. Cord BG analysis is typically performed only when an infant is depressed at birth, following a difficult delivery, or when there has been a nonreassuring fetal heart rate tracing. Thus, a large majority of infants in our cohort were missing BG data. Where available, the BG results were used to define whether the newborns endured fetal hypoxemia progressing to acidosis, a biomarker of HIE. Infants who died or required neonatal intensive care and for whom cord BG or neonatal BG results were unavailable were excluded from the study. In those cases, a definitive hypoxic etiology could not be confirmed without BG to measure fetal acidosis. We defined four mutually exclusive outcome groups [8,24]:

- **The healthy without BG group** (***N*** = 139, 512) included infants without adverse outcomes of labor or neurological injury for which clinicians did not request BG testing.
- **The healthy with BG group** (***N*** = 31, 734) had normal BG values: cord-gas pH ≥ 7.0 and base deficit BD *<* 10 mmol*/*L, or neonatal-gas BD *<* 10 mmol*/*L within the first two hours of birth. In addition, other neonatal markers were reassuring: (a) the Apgar score at five minutes from birth was ≥ 7, (b) no neonatal intensive care unit admission, (c) discharged home, (d) no death or hospital transfers in the neonatal period, (e) no neurological evidence of neonatal encephalopathy, (f) no intubation or chest compression during resuscitation, (g) no seizures were observed, and (h) therapeutic hypothermia was not required.
- **The acidosis group** (***N*** = 2, 636) was defined by cord-gas pH *<* 7.0 or BD ≥ 10 mmol*/*L, or neonatal gas BD ≥ 10 mmol*/*L within 2 hours from birth. Infants in this group did not have signs of neonatal encephalopathy during the first six hours of life.
- **The HIE group** (***N*** = 304) presented two mandatory criteria: acidosis as defined above, and an abnormal neurological examination during the first six hours of life confirmed by clinician chart review [8]. In addition, every infant in this group presented at least one of the following: (a) neurological abnormalities persisted beyond six hours after birth, (b) presence of neonatal seizures during the first 24 hours of life, or (c) therapeutic hypothermia was required.

#### 2.2.2 Clinical characteristics

The study cohort comprised 174,186 births in total. Their maternal, fetal, and intrapartum characteristics are summarized in Table 2. Labor duration, and consequently the number of CTG epochs per infant, increased steadily by group severity from the healthy to HIE groups. The rate of all Caesarean deliveries and Caesarean deliveries for fetal intolerance was greater in the HIE and acidosis groups than in the healthy groups. Finally, nulliparity and maternal fever were also more common in the HIE and acidosis groups than in the healthy groups [25]. For binary classification, the healthy with and without blood gas groups were treated as the healthy class, and the acidosis and HIE groups were treated as the pathological class. However, the results were evaluated for each subgroup separately.

**Table 2.** Clinical Characteristics and CTG Data Availability of the Study Population.

| Characteristic | Healthy without BG<br>($N = 139,512$) | Healthy with BG<br>($N = 31,734$) | Acidosis<br>($N = 2,636$) | HIE<br>($N = 304$) |
| --- | --- | --- | --- | --- |
| Gestational Age (weeks)<sup>a</sup> | 39.57 [38.86–40.43] | 39.86 [39.00–40.71] | 40.00 [39.00–40.71] | 40.00 [38.86–40.71] |
| Nulliparity (%) | 42.05% | 60.13% | 67.64% | 70.07% |
| Maternal fever (%) | 6.69% | 17.03% | 26.02% | 39.14% |
| Labor duration (hours)<sup>a</sup> | 8.92 [4.83–15.00] | 12.58 [6.68–20.33] | 13.67 [7.93–21.67] | 19.06 [10.23–27.85] |
| Valid 20-min epochs per infant<sup>a</sup> | 18 [9–32] | 27 [13–45] | 30 [14–50] | 43 [21–64] |
| Caesarean delivery (%) | 7.29% | 32.33% | 31.98% | 49.01% |
| Caesarean delivery for fetal intolerance (%) | 4.26% | 27.20% | 28.95% | 43.42% |

<sup>a</sup> Data presented as Median [Interquartile Range].

#### 2.2.3 Determination of labor onset

We identified labor onset computationally from the UA signals and clinical records. These data were assessed sequentially in 20-minute epochs. First, we identified the time at which regular contractions began in the UA signal. Then, labor onset was determined when one of the following was reported:

- rupture of membranes (ROM)
- cervical dilation greater than 4 cm
- any change in cervical effacement
- any change in cervical dilation

For ROM, labor onset was defined as the start of the evaluated 20-minute epoch. For the other three criteria, labor onset was defined as the time of the pelvic examination. If UA was not available, labor onset was based solely on the cervical-examination criteria. Finally, all cases that met none of these criteria were excluded from this study.

### 2.3 Signal processing and feature extraction

Following our previously described methodology [22,24], raw FHR and UA signals, sampled at 4 Hz, were preprocessed using a commercial FDA-cleared CTG analysis system (PeriCalm Patterns, PeriGen Inc.). During preprocessing, maternal-heart-rate interference, sensor dropout, and high-noise segments were removed [26,27]. FHR gaps shorter than 15 seconds were linearly interpolated, while longer gaps were excluded from analysis. Next, the signals were segmented into sequences of non-overlapping 20-minute epochs. The epochs were aligned with respect to the time of delivery. Thus, epoch *t* ∈ {1, 2*, . . .,* 216}, with *t* counting backwards from birth, indicates directly the lead time of each epoch; *t* = 1 indicates the epoch that started 20 minutes before delivery and covers up to the time of delivery, while *t* = 6 indicates the epoch that started 2 hours before delivery and spans up to 1 hour 40 minutes before delivery. This indexing notation allowed us to keep track of the lead time of the predictions when testing the classifiers and is used only to report results. In contrast, TLO was measured forward from the labor onset and computed for each epoch. Epochs with less than 80% valid signal coverage were excluded from the analysis. Across all infants, a total of 4,338,519 valid epochs were extracted and used in model training and continuous evaluation.

PeriCalm Patterns was also used to identify clinically relevant events from the CTG signals: FHR baseline, accelerations, decelerations, and uterine contractions [26,27]; PeriCalm Patterns further sub-classified each deceleration as [26]: *variable* decelerations fall rapidly from baseline, whereas the nadir of *late* decelerations follows the peak of the associated contraction. Baseline segments were present in every epoch, while accelerations, decelerations, and contractions were present in 63.76%, 39.80%, and 94.82% of epochs respectively; this missingness motivates the surrogate splits described in Section 2.5. We extracted a total of 40 morphological and fetal heart rate variability features, which were previously shown to be significantly associated with HIE [22]. The mathematical formulation of these features is detailed in our previous work [22,23]. Table 3 shows the complete list of features extracted from each CTG event type.

**Table 3.** Complete list of CTG features extracted from each event type.

| CTG event | Features extracted |
| --- | --- |
| Baseline (14) | *Morphological*: slope, dwell time, transitions to acceleration, transitions to deceleration. *Time domain*: mean FHR value, interval index. *Frequency domain*: low-frequency power, movement-frequency power, LF/(MF+HF) ratio. *Phase-rectified signal averaging*: deceleration reserve. *Non-linear*: approximate entropy ($r = 0.2$), sample entropy ($r = 0.1$), Hurst exponent, Lyapunov exponent. |
| Acceleration (3) | *Morphological*: event count, transitions to baseline. *Time domain*: mean FHR value. |
| Deceleration (20) | *Morphological*: event count, count of variable decelerations, count of late decelerations, count of decelerations that are neither variable nor late, area, depth, dwell time, transitions to acceleration, transitions to baseline. *Time domain*: mean FHR value, interval index. *Phase-rectified signal averaging*: deceleration reserve. *Non-linear*: approximate entropy ($r = 0.1, 0.2, 0.3$), sample entropy ($r = 0.2, 0.3$), correlation dimension, Hurst exponent, Lyapunov exponent. |
| Contraction (3) | *Morphological*: event count, dwell time, resting interval. |

The number of features per event type is given in parentheses. Mathematical formulations are given in [22].

### 2.4 Accounting for time from labor onset

To test our hypothesis that intrapartum classification should account for the evolution of FHR and UA properties in labor, we implemented two distinct structural modifications: (1) we trained models on progressively longer windows of *W* hours before delivery instead of the standard last-hour segment where *W* = 1 h (Section 2.7), and (2) we explicitly embedded TLO into the feature space. TLO quantifies the time spent from the beginning of labor; TLO = 0 indicates labor onset [24]. For every infant *i*, the feature vector ***x****_i,t_* was extracted from their *t* -th epoch. Thus, the time-aware feature vector was:

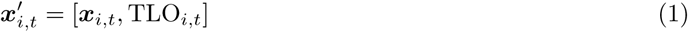

Appending TLO*_i,t_* as a feature allows the classifier to evaluate the CTG features conditionally: the classifier may learn to apply different pathological weights to an identical deceleration morphology depending on when in labor it occurs. We compare classifiers trained with and without this explicit index to isolate the performance gains obtained by a time-aware approach.

### 2.5 Classification framework

We formulated a binary classification task to discriminate infants at risk of severe acidosis or HIE (pathological) vs. healthy controls.

Classification was performed using Random Forest (RF) ensembles of 1,000 decision trees with surrogate splits. Surrogate splits are alternative splitting rules stored at each tree node, defined on predictors that are correlated with the primary splitting variable. These surrogates are triggered when the primary variable is missing. Since our features are extracted from each CTG event, surrogate splits enable the classifier to properly process all epochs even when some events may be missing from an epoch.

The dataset contained severe class imbalance (pathological cases represent only 1.7% of the cohort) and further imbalance between the four study subgroups. To address this, balancing was applied at the individual tree level using two settings that act together: observation weights determine the distribution of subgroups in the bootstrap sample, and the in-bag resampling fraction determines the size of the bootstrap sample. Each training epoch ***x****_i,t_* in the study subgroup *g* received a weight *w_i,t_* = *M/*(*G* · *M_g_*), where *G* = 4 is the number of subgroups, *M_g_* is the count of training observations in subgroup *g*, and *M* is the size of the training set. The weights of each subgroup add up to *M/G*, which means that each draw is equally likely to come from any subgroup. With minority class samples *M*_min_ = min*_g_*(*M_g_*), the in-bag fraction was set to *F* = *G* · *M*_min_*/M*, which gives a bootstrap sample size of *M*_min_ expected from each subgroup. Thus, each tree is trained on an approximately balanced sample of epochs from each subgroup, whose size is determined by the smallest subgroup (HIE). Different trees draw different subsets of the majority subgroups, allowing the full training set to contribute to the ensemble with no observations discarded.

During training, the RF treated each epoch as an independent observation. Per-infant aggregation and sequential testing were handled afterwards during the deployment simulation.

### 2.6 Cross-validation and threshold calibration

We evaluated the classifiers using repeated nested cross-validation. Usually, nesting is used to select model hyperparameters in the inner loop. In our study, RF hyperparameters were fixed and identical for every model. Instead, we used nested cross-validation solely to obtain prediction scores for the outer-training set from classifiers that had not been trained on them (validation predictions). These scores were later used to set the decision threshold *τ* for the classifier without touching the outer-test predictions.

All partitioning was performed at the infant level. This means that every epoch from a given infant was assigned to exactly one partition; no infant ever contributed epochs to more than one set within a given split. Sequential epochs from a single infant are highly correlated, so splitting epochs from any infant across the training and test sets would leak information and inflate the performance estimates. We stratified the partitions by outcome subgroups (healthy without BG, healthy with BG, acidosis, HIE), Caesarean delivery for fetal intolerance to labor, and nulliparity. Stratification ensures that the joint distribution of these variables remained stable across folds. The complete pipeline for model training, threshold calibration, and model testing is described in Algorithm 1.

**Algorithm 1.**
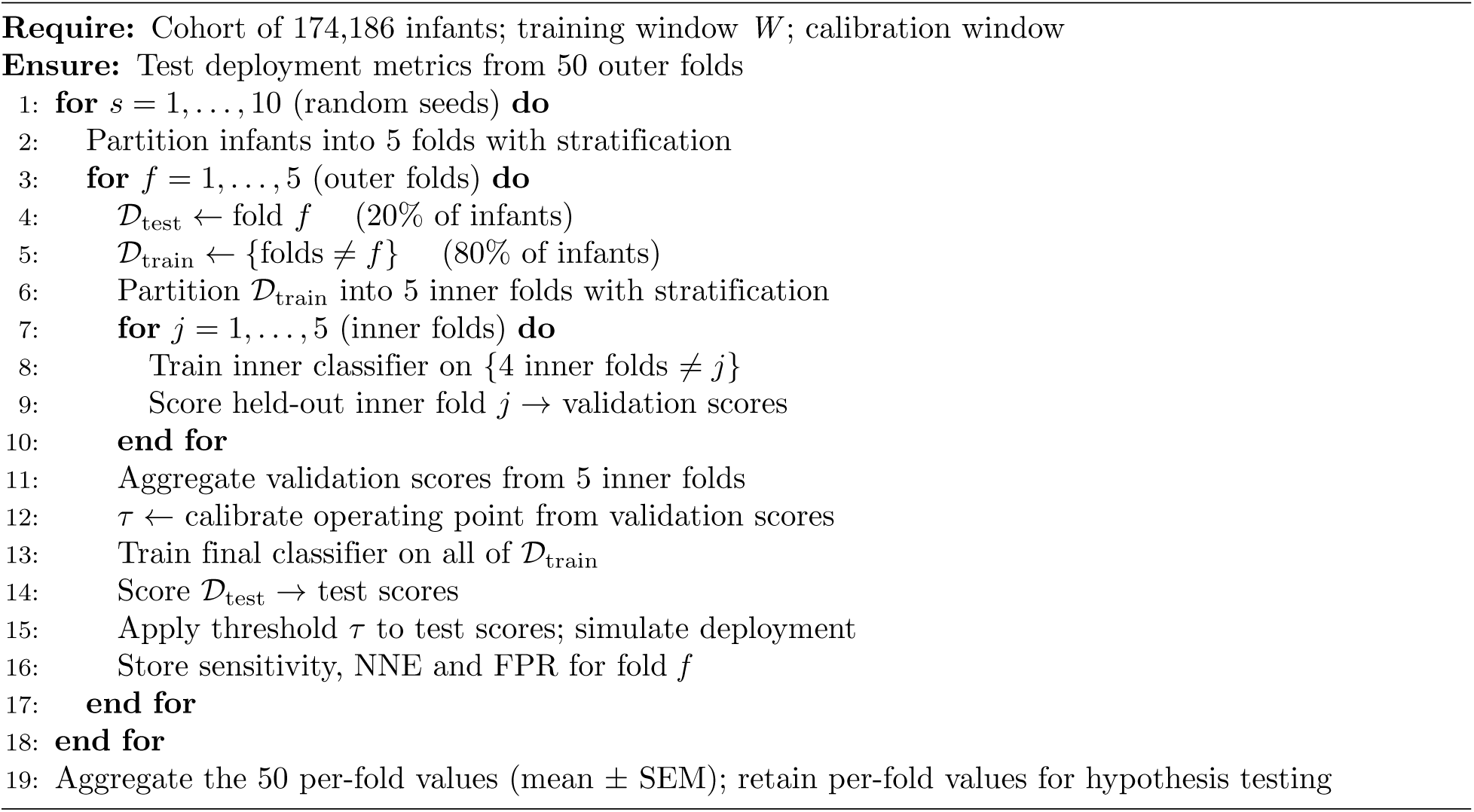
Repeated nested cross-validation and threshold selection.

For each infant *i* and their *t* -th epoch, the RF provides a score *P_i,t_* ∈ [0, 1] that the infant belongs to the pathological class. Thus, every infant receives a sequence of scores through all their epochs in labor. A standard classifier would assign the pathological label whenever *P* ≥ 0.5. This default is inappropriate here. Per-tree balancing makes the prediction scores approximate posteriors under a balanced class prior rather than the observed 1.7% prevalence of the cohort, so 0.5 carries no clinical meaning. More importantly, in intrapartum monitoring false positives translate into unnecessary clinical interventions, so the operating point must be carefully controlled such that the alarm burden is clinically tolerable. In this study, we set the false positive rate (FPR) to not be greater than 15% for all the models during calibration. This served two purposes: (1) it held the alarm burden constant across all comparisons, so that improvements in sensitivity are directly comparable, and (2) it also matched the operating point at which prior work reported sensitivity [10,13], allowing our values to be compared against published results. In this cohort a 15% FPR corresponds to approximately 25,687 healthy infants receiving at least one alarm; the alarm burden per true detection is quantified by the number needed to evaluate (NNE) in Section 2.8.

The operating point was determined from the validation scores of Algorithm 1. Because clinical interventions occur at the infant level, the scores of each infant were aggregated to a single value before percentile selection:

1. Restrict the validation scores to a calibration window, which depends on the experiment and is defined in Section 2.7.
2. For every outer-training infant, take the maximum score *P ^∗^* across all of their epochs within the calibration window.
3. Retain only the infant-level maxima of the healthy class, such that *τ* is defined by the alarm burden we aim to control.
4. Set *τ* to the 85th percentile of *P ^∗^*, giving FPR ≤ 15%.

A single *τ* is computed per experiment per outer fold and is applied to the test predictions of that fold. No recalibration on the hold-out set occurs. We pooled the validation scores across the five inner classifiers before percentile selection. These classifiers differ only by the subset of the outer-training set they were fitted to; they share an identical feature space, hyperparameters and balancing scheme. Since threshold selection depends on the healthy class, the largest in the cohort, it is reasonable to expect that the distributions of classification scores across two random subsets of the same class are not different. Thus, the operating point defined on a subset of the training set should yield a comparable FPR in the hold-out cross-validation set.

### 2.7 Evaluation frameworks

In Section 2.6 we defined the 10 × 5 cross-validation approach and *τ* calibration. Every experiment applies that same procedure and differs only in three time windows:

- the **training window**: where the classifier sees data;
- the **calibration window**: where *τ* is set (Section 2.6);
- the **evaluation window**: where test performance is measured.

Except for the idealized last-hour test defined below, the evaluation window ran from labor onset to epoch *t* = 3, which spans from 1 hour to 40 minutes before delivery. Discarding the last two epochs before delivery implies that any clinical decision supported by our classifier is actionable. Thus, we define 40 minutes before delivery as the *actionable deadline*.

We trained two types of classifiers: (1) time-invariant (TI), which map the feature space to scores without knowledge of when in labor the epoch occurred, and (2) time-aware (TA), which received TLO as an additional input, conditioning the mapping on timing with respect to labor onset. Table 4 specifies the three approaches being compared.

**Table 4.** Training, Calibration, and Test Specifications of the Three Approaches.

| Approach | Training window | Calibration window | Evaluation window |
| --- | --- | --- | --- |
| Standard (TI, 1 h) | Last hour | Last hour | Last hour and full labor |
| Extended (TI, $W$) | $W$ | Full labor | Full labor |
| Time-aware (TA, $W$) | $W$ | Full labor | Full labor |

Full labor: labor onset to actionable deadline. $W$ : Varying size training window.

#### 2.7.1 Standard approach

The classifier was trained and *τ* was calibrated using only the final hour of labor (*t* ∈ {1, 2, 3}). Then it was evaluated under two lenses: the **idealized standard test**, restricted to the final hour, and the **deployment test**, in which *τ* drives a sequential alarm simulation across the entire labor up to the actionable deadline *t* ≥ 3. The standard test mirrors published last-hour methodology, while the deployment test reveals what happens when such a model is operated continuously. This experiment compared the behavior of the 1-hour TI classifier, with last hour *τ* calibration, in two different evaluation windows. We also filtered the test infants to those that had at least one epoch available in *t* ∈ {1, 2, 3} and in *t* ≥ 3; the same infant must have data available in both evaluation windows. This filtering was carried only for this evaluation and enabled the comparison across different evaluation windows under the same subset of infants. The two evaluation windows share epoch *t* = 3. Thus, infants whose only available epoch is *t* = 3 have identical scores under both lenses, making the comparison conservative.

#### 2.7.2 Extended-window approach

The classifier was trained on a longer historical window *W* ∈ {1, 3, 6, 9, 12, 18, 24, 36, 48, 72} h before delivery, and *τ* was calibrated on infant-level maxima across all *t* ≥ 3. In this experiment, the *W* = 1 h configuration uses the same training data as the standard approach but calibrates *τ* over the full labor, beyond the final hour. Also, this approach is evaluated on the unrestricted test set. Thus, the two report different performance for the same trained classifier.

#### 2.7.3 Time-aware approach

The classifier is trained using a window *W* and the TLO feature appended to the predictor set. This experiment isolates the additional benefit of explicit temporal indexing, while keeping the training-window length and calibration approach constant. The calibration and evaluation windows were identical to the extended-window experiment, such that the two differ only in the feature set and the effect of TLO is not confounded with a change in calibration.

### 2.8 Performance metrics

Continuous deployment was simulated by walking through the list of classifier outputs per infant, sequentially starting from labor onset. The first epoch with *P_i,t_* ≥ *τ* triggers an alarm. Epochs after the actionable deadline *t* ∈ {1, 2}, are ignored and cannot trigger an alarm; an alarm fired later than this deadline would carry no clinical utility. Model performance was summarized by the following metrics:

- **Cumulative true positive rate (sensitivity):** the proportion of pathological cases, stratified by subgroup (acidosis or HIE), that received at least one alarm at any point in their labor trajectory before the deadline. We report this both at the deadline itself (*t* = 3) and at the earlier landmarks of 3 h (*t* = 10) and 6 h (*t* = 19) before delivery to quantify early-detections.
- **Total false positive rate:** the proportion of healthy cases that triggered at least one alarm at any point at *t* ≥ 3; this is the number of false alarms that could trigger interventions.
- **Number needed to evaluate (NNE):** the ratio of all infants detected as pathological divided by the number of true HIE or acidosis detections. This is the inverse of the precision.
- **Area under the receiver-operating characteristics curve (AUROC)**: the scores of each infant were aggregated as their maximum score through the complete deployment window. Thus, each infant was associated with only one classification score, the most severe score through labor. Then, the AUROC was computed from the trade-off between sensitivity and 1-specificity as the decision threshold varied from 0 to 1.

Reporting these metrics up to the actionable deadline ensures that every alarm that was counted in the sensitivity score is one that, in principle, could have prompted a clinical intervention.

### 2.9 Optimal-window selection

The optimal training window was selected using a step-down procedure on the early-detection rate of the HIE subgroup, the clinical target of this study; severe acidosis is reported as a secondary outcome. Selecting on the pooled performance of the pathological class would have biased the criterion toward acidosis, which is 8.7 times more prevalent. This choice was specified before the comparisons were run.

The step-down procedure for window length selection allowed us to identify the shortest training window that achieves a performance not statistically different from the best performer. First, we applied a Friedman omnibus test to compare the HIE detection at 3 h (*t* = 10) before delivery across all training windows *W*. If non-significant differences were found, all windows were declared equivalent and the shortest was selected. Otherwise, we defined *W* _peak_ as the window with the highest mean HIE detection at 3 h. Windows lying within one standard error of the maximum were considered ties; these were resolved using the HIE detection at 6 h (*t* = 19). With this selection, we tested whether shorter windows had significantly different performances from *W* _peak_ when tested under pairwise comparisons. The one-sided Wilcoxon signed-rank test was applied as: H_1_ : *W*_peak_ outperforms the shorter windows at 3 h, and at 6 h before delivery.

Holm-Bonferroni correction was applied to all 2 · |{*W* : *W < W*_peak_}| tests. A shorter window was declared statistically indistinct from *W* _peak_ only if both adjusted *p* ≥ 0.05, for the comparisons at 3 h and at 6 h. The optimal *W* ^*^ was the shortest window not statistically different from *W* _peak_; if none qualified, then *W ^∗^* = *W*_peak_.

### 2.10 Model interpretability and feature importance

Feature importance was estimated post-hoc by permutation testing on the test predictions of each fold. The loss function was the drop in infant-level partial AUROC (pAUROC) over the FPR range 0–15%.

Predictions were aggregated at the infant level by taking the maximum classification score within the relevant time window, as in Section 2.6, so that each infant contributes a single observation and the importance estimate is aligned with the deployment evaluation. Each permutation was carried out on a balanced bootstrap subset of the test infants. In every shuffle, *M* _min_ infants – the number of infants from the

HIE group – were drawn with replacement from each of the four subgroups. For each feature, the cycle of bootstrap resampling, permutation, model testing, infant-level aggregation, and pAUROC computation was repeated 10 times and the resulting drops averaged. The importance of a feature was defined as the ratio between the drop in pAUROC due to its permutation and the original pAUROC of the model, averaged across the 10 × 5 folds. Per-fold values were retained for significance testing.

- **Global importance** was computed by aggregating scores over the deployment window. This allowed us to assess which features had the largest impact on the discriminability of infants throughout all labor. When comparing two different models, feature importance estimates were compared with the Wilcoxon signed-rank test and Holm-Bonferroni correction.
- **Time-resolved importance** was computed for the TA classifier only, applying the same procedure independently within each one-hour TLO bin. This procedure generated per-feature importance trajectories across labor. Because TLO is almost constant within a one-hour bin, permuting it would not perturb the model input and it was therefore held fixed. The analysis was restricted to

TLO ≤ 10 h, approximately the median labor duration of the healthy class, whose size determines the false-positive axis of the pAUROC. Here, we report the time-resolved importance for the features with the highest global importance for the TA classifier. Since the expected permutation importance of an irrelevant feature is zero – disrupting a feature cannot improve predictive performance [28] – importance estimates below zero were deemed irrelevant and ignored. These bins were masked as a descriptive visual aid and not a formal inferential claim.

## 3 Results

### 3.1 The deployment evaluation of the standard approach

Table 5 compares the standard 1-hour TI classifier, evaluated under its idealized last-hour test, against continuous deployment. In the restricted test set, AUROC and sensitivity rose for both subgroups: AUROC had a larger increase for HIE (0.709 to 0.779) than for acidosis (0.719 to 0.726), and sensitivity increased from 46.8% to 58.5% for HIE and from 46.8% to 48.8% for acidosis. All these differences were statistically significant (*p <* 0.001). The two pathological subgroups diverged in their NNE: the acidosis NNE increased from 22.4 to 23.4, one additional infant evaluated per true detection (4.3%), whereas the HIE NNE decreased from 193.8 to 166.9 (13.9%). The FPR rose from 14.9% to 16.2%, because *τ* was calibrated only in the last hour of labor and then applied across the complete labor.

**Table 5.** Standard 1-Hour TI Test vs. Continuous Deployment.

| Metric | Standard Test<br>(Final Hour) | Deployment<br>(Continuous) | Adjusted<br>*p*-value |
| --- | --- | --- | --- |
| AUROC (Acidosis) | $0.719 \pm 0.002$ | $0.726 \pm 0.002$ | **0.0009** |
| AUROC (HIE) | $0.709 \pm 0.006$ | $0.779 \pm 0.005$ | **< 0.0001** |
| Sensitivity (Acidosis) | $46.8\% \pm 0.3\%$ | $48.8\% \pm 0.3\%$ | **< 0.0001** |
| Sensitivity (HIE) | $46.8\% \pm 1.0\%$ | $58.5\% \pm 0.9\%$ | **< 0.0001** |
| NNE (Acidosis) | $22.4 \pm 0.2$ | $23.4 \pm 0.2$ | **< 0.0001** |
| NNE (HIE) | $193.8 \pm 4.5$ | $166.9 \pm 2.9$ | **< 0.0001** |
| False Positive Rate | $14.9\% \pm 0.0\%$ | $16.2\% \pm 0.1\%$ | **< 0.0001** |

Analysis performed on a restricted test set (Section 2.7). Data is mean ± SEM. *p*-values from two-sided Wilcoxon signed-rank test and Holm-Bonferroni correction.

The two evaluation windows do not completely overlap; the standard test includes the final 40 minutes of labor, which the deployment lens discards. Even so, the deployment test detects more HIE cases despite not having access to the epochs closest to birth.

### 3.2 Training window expansion

Fig 2 shows model performance as the training window *W* grew, and S1 Table reports all values. The Friedman test rejected the equivalence of early HIE detection across windows at both 3 h and 6 h before delivery (both *p <* 0.0001). HIE sensitivity at these landmarks was highest at *W* = 72 h (37.1% at 3 h and 25.2% at 6 h), against 32.0% and 16.6% for *W* = 1 h. Five windows fell within one SEM of the maximum at 3 h (*W* ∈ {18, 24, 36, 48, 72}), so the tie was resolved with the HIE sensitivity at 6 h defining *W*_peak_ = 72 h.

**Fig 2.**
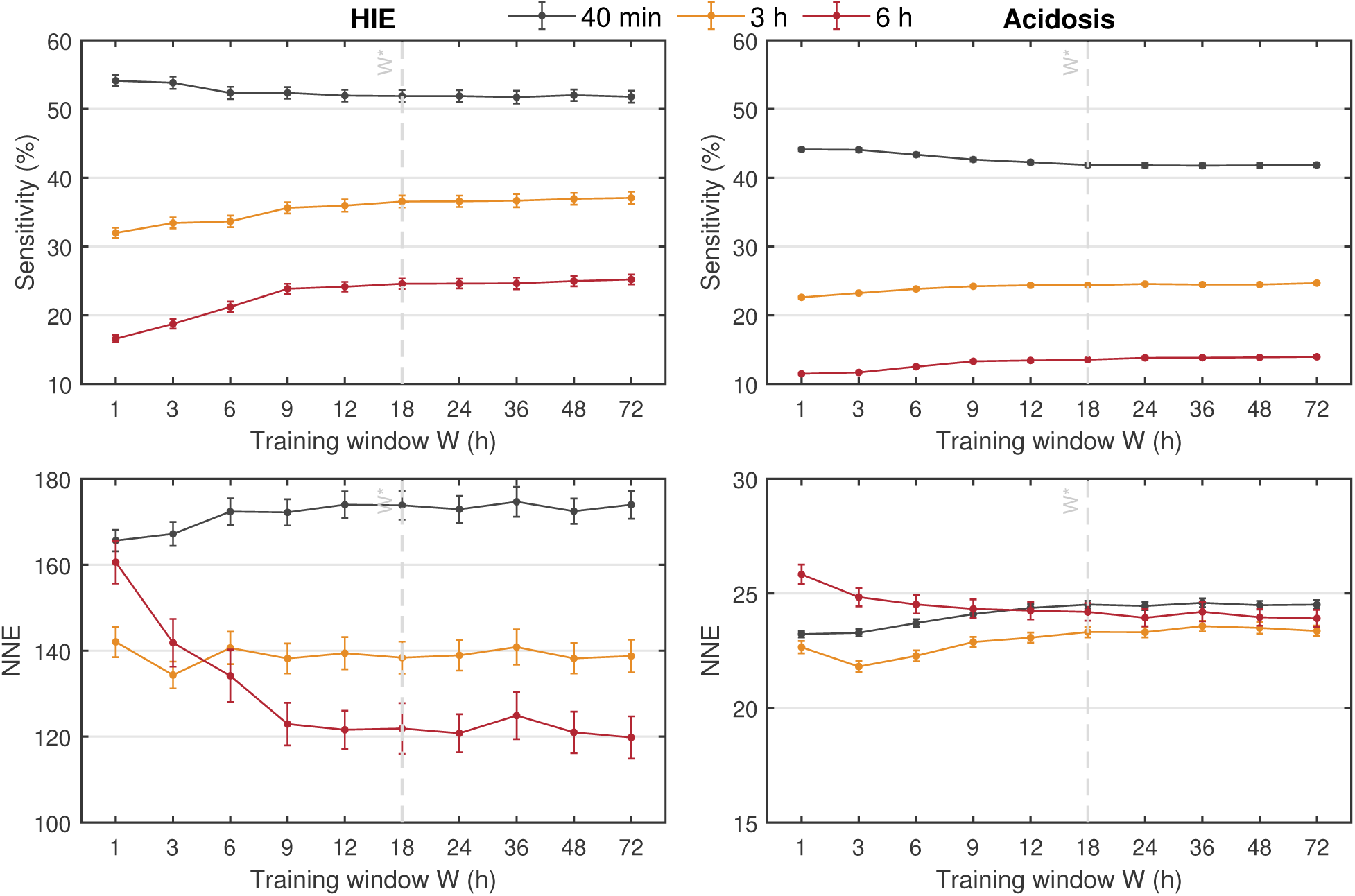
Optimal-window selection. Sensitivity and NNE obtained in the HIE and Acidosis groups as a function of the size of the training window *W*. The performance metrics are reported at three times before delivery: (1) at the actionable deadline of 40 minutes before delivery, (2) at 3 h before delivery, and (3) at 6 h before delivery. Data is presented as mean ± SEM.

The step-down procedure selected *W ^∗^*= 18 h, which was the shortest window whose HIE sensitivity was not significantly lower than *W* _peak_ at 3 h (*p* = 0.48) and 6 h before delivery (*p* = 0.37); the next shorter window *W* = 12 h failed at 6 h (*p* = 0.0268), and *W* = 9 h failed at both landmarks (*p* = 0.0406 and *p* = 0.0162).

Fig 2 shows that the performance at 40 min, 3 h, and 6 h moved in different directions as *W* increased. HIE sensitivity at 3 h and 6 h rose consistently up to approximately *W* = 18 h and plateaued thereafter, while HIE sensitivity at 40 min before delivery declined slightly and then plateaued. The HIE NNE mirrored this: it fell substantially at 6 h and rose at 40 min. In the acidosis group the same behavior was observed but at a much smaller magnitude, and the acidosis NNE varied by less than 2 across the entire sweep. While longer training windows shifted detection earlier in labor, the deployment false positive rate remained stable within the 15.0–15.1% range, as reported by S1 Table.

### 3.3 Best TI classifier and TLO indexing

Fig 3 shows the cumulative sensitivity curves for the 1-hour TI, 18-hour TI, and 18-hour TA classifiers, and S2 Table reports the corresponding performance metrics at the landmark times with pairwise comparisons (computed on the full test set, unlike Table 5). The FPR remained at the 15% target for all three classifiers, and no pairwise difference in FPR was significant.

**Fig 3.**
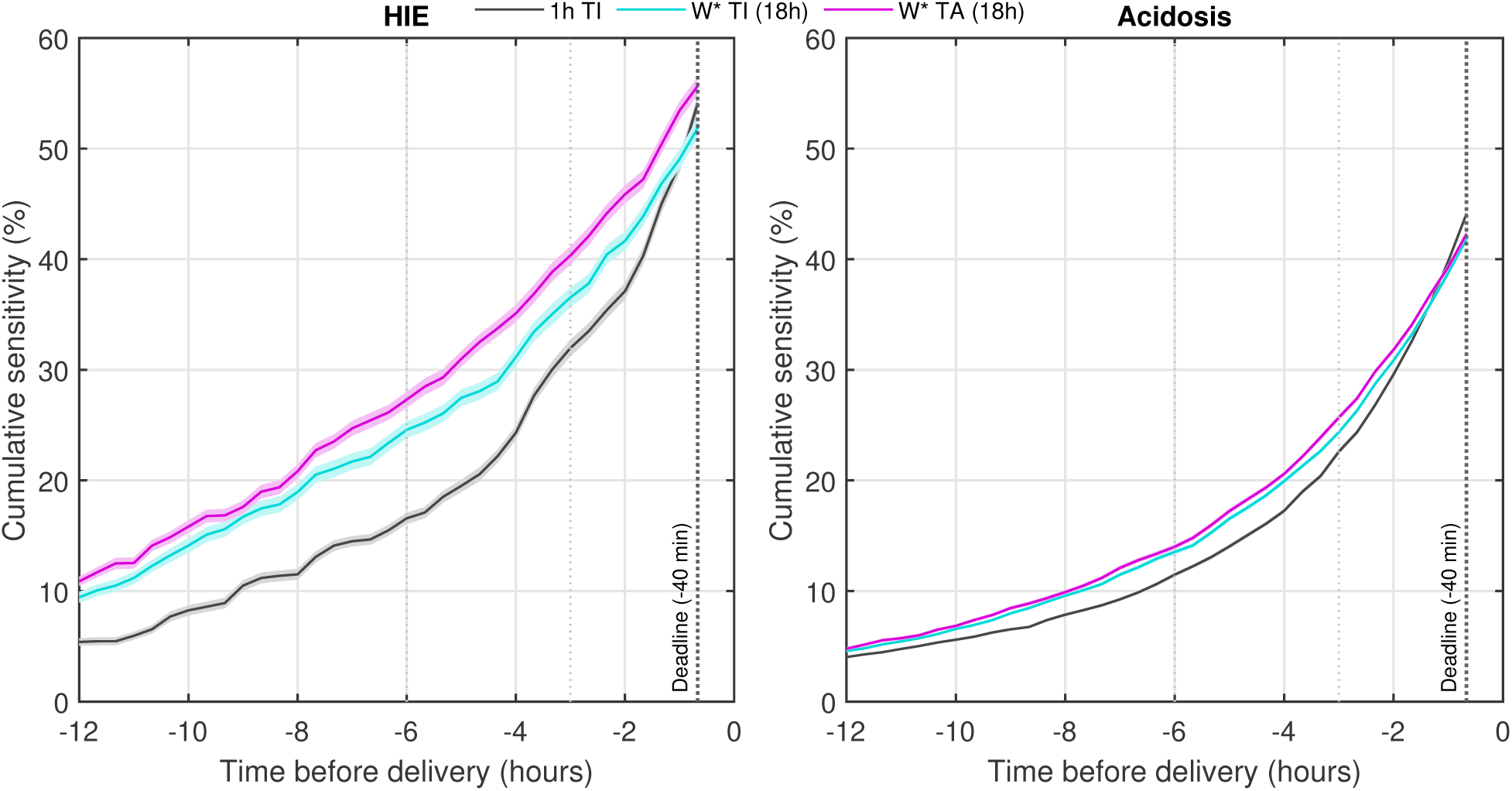
Cumulative sensitivity. The proportion of detections obtained in the HIE and acidosis groups using the three models: (1) TI classifier with *W* = 1 h, (2) TI classifier with *W ^∗^* = 18 h, and (3) TA classifier with *W ^∗^* = 18 h. All sensitivities are reported continuously in the last 12 h of labor up to the actionable deadline to quantify lead time. Data is presented as mean ± SEM (shaded area).

For HIE, expanding from *W* = 1 h to *W* ^*^ increased sensitivity by 4.6% at 3 h and 8.0% at 6 h (both *p <* 0.001), and decreased it by 2.2% at the actionable deadline (*p <* 0.01). Adding TLO increased HIE sensitivity further at all three landmarks: 3.8% at the deadline, 3.8% at 3 h, and 2.7% at 6 h (all *p <* 0.001). The 18-hour TA classifier achieved sensitivities of 27.3%, 40.3%, and 55.7% at 6 h, 3 h, and 40 min before delivery respectively. Thus, relative to the 1-hour TI classifier, the 18-hour TA classifier detected 8.4% more HIE cases at 3 h and 10.7% at 6 h (both *p <* 0.001), and its sensitivity at the deadline was no longer reduced (+1.6%, *p* = 0.09). Also, the 18-hour TA classifier had the lowest HIE NNE at all landmarks (S2 Table): 161.5 at the deadline, 128.9 at 3 h, and 111.1 at 6 h, against 165.6, 142.0, and 160.6 for the 1-hour TI classifier. These improvements were significant at 3 h (−13.1, *p <* 0.001) and at 6 h (−49.5, *p <* 0.001) but not at the actionable deadline (−4.1, *p* = 0.14).

For acidosis, using *W* ^*^ increased sensitivity by 1.8% at 3 h and 2.1% at 6 h, and decreased it by 2.3% at the deadline (all *p <* 0.001). Adding TLO increased acidosis sensitivity further by 1.3% at 3 h (*p <* 0.001), with no significant change at 6 h (+0.5%, *p* = 0.06) or at the deadline (+0.4%, *p* = 0.27). Thus, both 18-hour classifiers detected fewer acidosis cases at the actionable deadline.

The same divergence was observed in the test AUROC, which is threshold-free. Relative to the 1-hour classifier, both 18-hour classifiers had a lower acidosis AUROC (0.699 to 0.690 and 0.690, both *p <* 0.001) and a higher HIE AUROC (0.766 to 0.774, *p <* 0.01; and to 0.790, *p <* 0.001). The 18-hour TA classifier had the highest HIE AUROC, 0.016 higher than the 18-hour TI classifier (*p <* 0.001), while their acidosis AUROC did not differ (*p* = 1.00).

Finally, Fig 4 shows the distribution of alerts obtained with the 18-hour TA classifier for the healthy without BG, the healthy with BG, the acidosis, and the HIE groups. These alerts represent the false positives in the healthy groups, and the true positives (sensitivity) in the pathological groups. The distributions show that at the three time landmarks – 6 h, 3 h, and 40 min before delivery – the alert rates had a wider distribution for the minority groups, corresponding to the pathological class, than for the majority groups, corresponding to the healthy class. Also, at all times, the alert rate was lower in the healthy without BG group than in the healthy with BG group; the healthy without BG group was over four times larger than the healthy with BG group and therefore drove down the total FPR.

**Fig 4.**
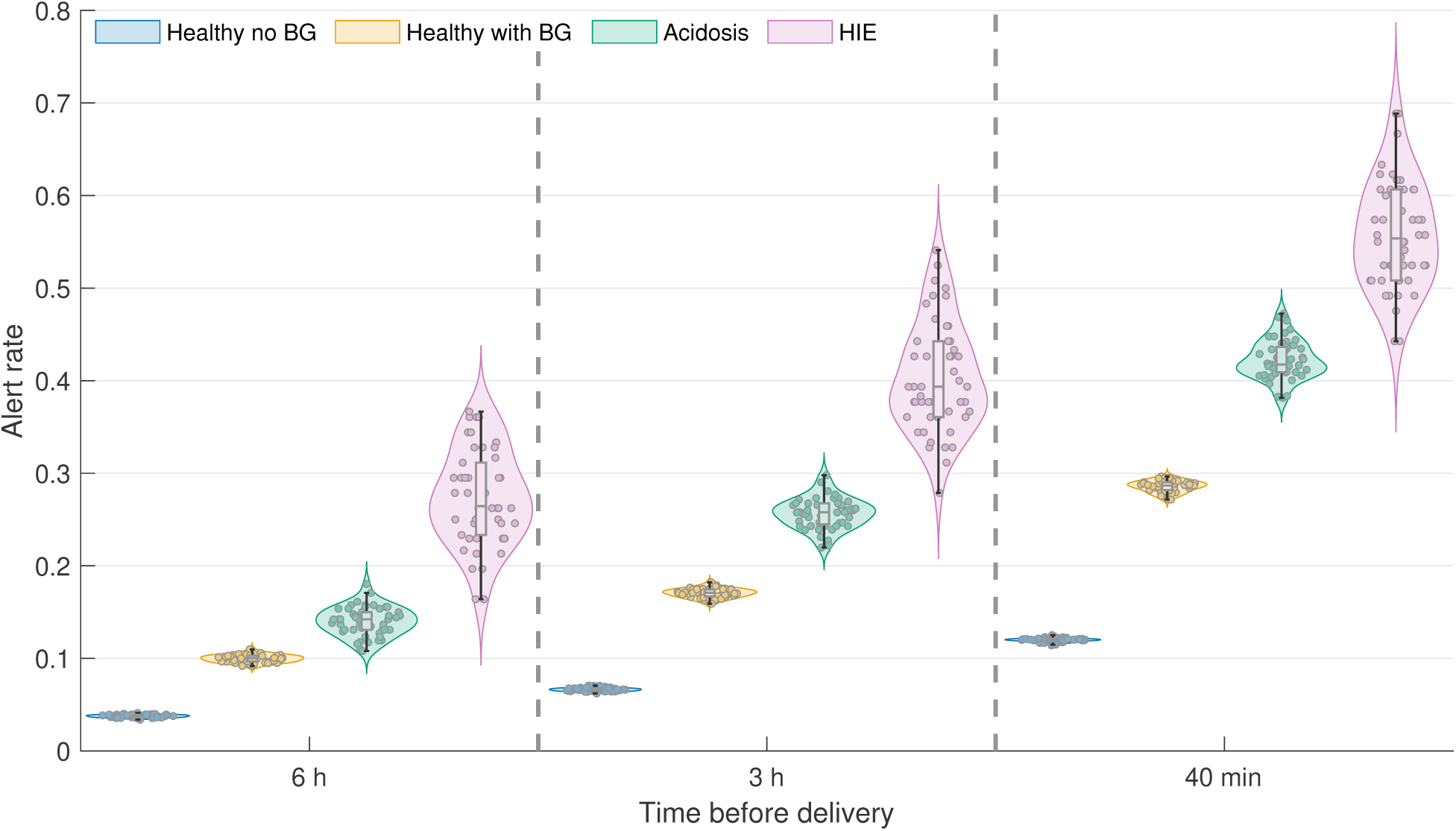
**Distribution of the alert rates for all groups in the cohort:** at 6 h, 3 h, and 40 min before delivery shown as violin plots. The distributions are plotted for the healthy without BG, the healthy with BG, the acidosis, and the HIE groups separately.

### 3.4 Feature importance analysis

S3 Table compares the TI global permutation importance for *W* = 1 and *W ^∗^* = 18. Only the mean value of the FHR baseline differed significantly: its importance almost doubled when the *W* was extended (5.08 to 9.65; medium effect, *r* = 0.47, *p* = 0.0318). No other feature had its importance change significantly. Thus, extending the training window made the model more focused on the FHR baseline without displacing the remaining features.

Table 6 reports the global permutation importance for the top 10 features of the 18 h TA classifier, and the corresponding importance estimates for the same features with the 18 h TI classifier. These results show that across all labor, the two features with the largest importance for the TA classifier were the mean value of the FHR baseline, and the time from labor onset. Permuting these features resulted in the pAUROC dropping by 6.03% and 4.93% respectively. This also indicates that in the TA classifier, TLO is almost as important as the most important CTG feature. Moreover, when compared to the TI classifier, the importance of most FHR features in Table 6 dropped once TLO was included. These differences were nominally significant for the baseline mean FHR, the deceleration dwell time and the deceleration interval index (*p* = 0.0025, 0.0254, 0.0473) but did not survive Holm-Bonferroni correction across all 40 features (Table 6). Thus, they are consistent with, but do not establish, the TA classifier reallocating its focus away from some of these CTG features towards TLO.

**Table 6.** Top 10 Features: Global Importance for the 18-hour TI and TA Classifiers.

| CTG Event | Feature | Permutation Importance |  | Mean difference | p-raw | p-adjusted |
| --- | --- | --- | --- | --- | --- | --- |
|  |  | 18 h TI | 18 h TA |  |  |  |
| Baseline | Mean FHR value | 9.65 ($\pm 1.02$) | 6.03 ($\pm 0.97$) | -3.62 | 0.0025 | 0.0990 |
| – | Time from labor onset | – | 4.93 ($\pm 1.16$) | – | – | – |
| Baseline | Dwell time | 1.27 ($\pm 0.33$) | 1.69 ($\pm 0.29$) | +0.42 | 0.3566 | 1.0000 |
| Deceleration | Dwell time | 2.51 ($\pm 0.39$) | 1.27 ($\pm 0.33$) | -1.24 | 0.0254 | 0.8902 |
| Deceleration | Interval index | 1.53 ($\pm 0.27$) | 1.05 ($\pm 0.18$) | -0.48 | 0.0473 | 1.0000 |
| Baseline | Transition to acceleration | 1.10 ($\pm 0.42$) | 0.47 ($\pm 0.44$) | -0.63 | 0.4371 | 1.0000 |
| Contraction | Count | 0.41 ($\pm 0.15$) | 0.33 ($\pm 0.18$) | -0.08 | 0.3771 | 1.0000 |
| Baseline | Lyapunov exponent | 0.19 ($\pm 0.23$) | 0.25 ($\pm 0.20$) | +0.06 | 0.7537 | 1.0000 |
| Deceleration | Count – not late nor variable | 0.11 ($\pm 0.24$) | 0.16 ($\pm 0.20$) | +0.05 | 0.8356 | 1.0000 |
| Deceleration | Area | 0.40 ($\pm 0.21$) | 0.02 ($\pm 0.21$) | -0.38 | 0.2184 | 1.0000 |

Data are mean ( ± SEM). Features are sorted from highest to lowest importance for the 18 h TA classifier; this classifier determined the top 10 being reported here. Differences were evaluated with Wilcoxon signed-rank test and $p$ -values were adjusted with Holm-Bonferroni correction across all 40 CTG features.

Finally, Fig 5 shows the time-resolved importance of the same features reported in Table 6, with the exception of TLO. The mean FHR baseline had the highest importance of any feature across all evaluated time bins, followed by the number of transitions between baseline and acceleration segments. The bin at which each feature reached its maximum was different. Features peaking in early labor (TLO ≤ 4 h) included the mean FHR baseline, the baseline dwell time, the deceleration count (not late nor variable), and the deceleration area. In contrast, features peaking later (TLO ≥ 5 h) included the deceleration dwell time, the deceleration interval index, the baseline to acceleration transitions, the contraction count, and the baseline Lyapunov exponent. Therefore, the importance of individual features varied with timing in labor.

**Fig 5.**
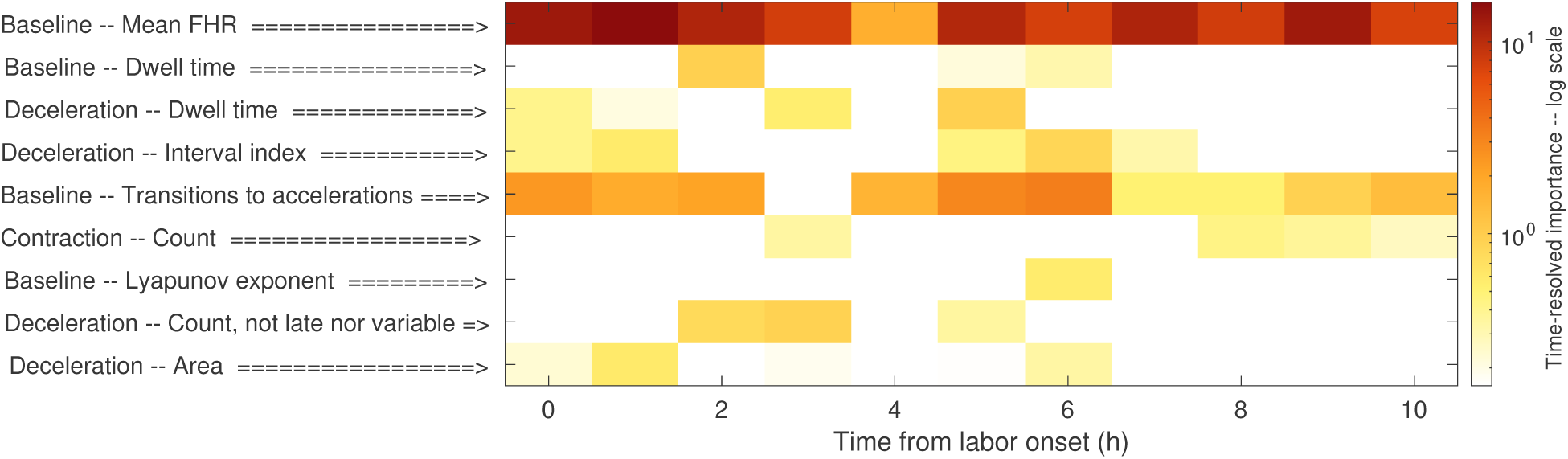
**Feature importance across labor (18-hour TA classifier):** for the CTG features displayed in Table 6, sorted in the same order. Importance is reported across the first 10 h of labor in log scale.

## 4 Discussion

In this study, we assessed whether the last hour before delivery is the appropriate window to train and evaluate classifiers aimed at the early identification of fetuses at risk of intrapartum HIE. Our results demonstrated that the final hour of labor is not representative of the behavior of the CTG signal throughout labor for HIE early-risk assessment. HIE shows discriminative signs much earlier than the final hour, and expanding the time window used to train a classifier increases significantly the number of HIE cases that may be identified early enough to prompt expert clinical evaluation. Indexing labor progression as TLO increased HIE sensitivity further, with the joint approach improving HIE sensitivity significantly throughout labor. Earlier detections enable clinical action, which is not possible with end-of-labor assessments. Thus, intrapartum decision support should evaluate labor as a continuous and time-aware process to improve the early identification of fetuses at increased risk of HIE.

### 4.1 The last hour is the wrong window for HIE assessment

Identifying fetuses at risk of acidosis and HIE in labor requires careful consideration of when the relevant signs appear. Acidosis generally develops before HIE, so it has an earlier impact on FHR characteristics. Uterine contractions intermittently reduce uteroplacental perfusion in proportion to their intensity [2], which reduces gas exchange at the placenta. Acidosis measured at birth is the cumulative result of this repeated stress. If acidosis persist – and their compensatory mechanisms fail – the fetus will sustain myocardial depression and impaired contractility, leading to fetal hypotension, cerebral hypoperfusion, and eventually neuronal energy failure as the onset of HIE [4,29]. In near-term fetal sheep subjected to intermittent, complete umbilical-cord occlusions at a period comparable to contractions in labor, arterial pH fell from 7.36 to 7.10 within 30 min and to 6.98 by half the occlusion series (∼1 h 22 min). In contrast, severe hypotension was not reached until a mean of 2 h 44 min. Moreover, three of eleven control fetuses tolerated 4 h of occlusions without ever developing severe hypotension, while the complete control group reached a mean pH of 6.91 [29]. Thus, severe acidosis can occur without the hemodynamic decompensation that precedes neurological injury. These experiments also indicate that a fetus progressing towards HIE is on a longer trajectory than one progressing towards acidosis alone, so its antecedents extend further back into labor. A classifier trained only on the last hour is trained on the physiology of acute compromise, dominated by contractions in the expulsive phase of labor, and can detect HIE only insofar as the infants progressing towards HIE show acute end-of-labor patterns. The studies in Table 1 justify the last-hour window by proximity to the outcome measurement or by following prior convention [10–13]. That rationale is sensible for acidosis, whose presence at birth indicates an onset during the final hours of labor. However, it does not transfer to an outcome whose antecedents begin much earlier.

The deployment test in Table 5 indicated that discriminating signs of HIE occur earlier than the last hour. This test used the same 1-hour TI classifier, in the same subgroup of test infants, with the same last-hour calibration, only shifting the evaluation window. Evaluating the model across labor significantly increased the HIE AUROC by 0.07 and sensitivity by 11.7%, even though the deployment test discarded the data closest to delivery. The same shift in the evaluation window only slightly increased the acidosis sensitivity by 2%. Importantly, the FPR rose from 14.9% to 16.2%, indicating that classifiers must calibrate their operating point over the window in which the model will actually run to ensure FPR stability. Moreover, the increase in false positives tells different stories for the acidosis and HIE groups when assessed in the context of the NNE. Even with more false positives, the HIE NNE dropped by 13.9% while it rose by 4.3% for acidosis. The gains in HIE may be due to the presence of its discriminating signs earlier in labor, whereas the gains in acidosis are due to the classifier looking at more epochs overall and not necessarily due to it finding earlier discriminating signs. If signs of HIE occur much earlier in labor than the final hour, then these classifiers can never learn other early discriminating signs of HIE besides those that are similar to the development of acidosis at the end of labor.

Training TI classifiers on longer windows improved the early identification of fetuses at risk of HIE. While expanding the training window likely introduces label noise into the classifier – early epochs of an acidosis or HIE case carry the label of the outcome of labor even when the fetus may have been healthy at labor onset – classification performance did not fall uniformly across outcomes and labor. Instead, it significantly improved early HIE sensitivity in particular. When expanding the training window from *W* = 1 h to *W ^∗^* = 18 h, acidosis AUROC fell (0.699 to 0.690, *p <* 0.001) while HIE AUROC rose (0.766 to 0.774, *p <* 0.01), and early HIE sensitivity improved in 8.0% at 6 h and 4.6% at 3 h before delivery (both *p <* 0.001). This improvement resulted only from expanding *W* for the TI classifier, without any information about TLO or labor duration. This gain in early sensitivity came at the cost of reduced sensitivity at the actionable deadline, which fell by 2.2% for HIE and 2.3% for acidosis. Expanding the training window redistributed the discriminative information away from acute end-of-labor deterioration towards the slower and progressive deterioration that precedes neurological injury. This interpretation relies on the aggregate performance of the TI models, and not on the physiology of individual tracings. Also, the permutation-importance results corroborate this only weakly: the importance of the mean FHR baseline nearly doubled from *W* = 1 to *W ^∗^* = 18, although no other significant changes in feature importance were observed (S3 Table).

For *W >* 18, early HIE sensitivity plateaued, which may be due to the distribution of labor duration. The median labor in the HIE group lasted 19.06 h (Table 2), so the last 18 h cover most of labor in these cases. Thus, the 72 h of data that is available for the longest records is not required to train a classifier: roughly the median labor duration of the target class suffices. The 18-hour requirement only pertains to the training data and not deployment: our classifier processed one 20-minute epoch at a time, so no history buffer is needed during continuous monitoring.

### 4.2 The advantage of TLO

Adding TLO to the 18-hour TI classifier improved early HIE sensitivity further: it rose by 3.8% at the actionable deadline, 3.8% at 3 h, and 2.7% at 6 h before delivery (all *p <* 0.001). NNE also fell significantly at the deadline and at 3 h but not at 6 h. Thus, more HIE cases were identified by the TA classifier without a proportional increase in the number of alarms, and FPR remained stable. Moreover, the improvement in discriminability was specific to HIE, for which AUROC increased by 0.016 with no significant difference for acidosis. Only the sensitivity at 3 h improved significantly for the acidosis group, and it did not change significantly at the other landmarks.

Moreover, adding TLO recovered HIE sensitivity at the deadline, which was indistinguishable from the sensitivity of the 1-hour TI classifier (+1.6, *p* = 0.09). This indicates that the trade-off observed when extending the training window may not be intrinsic to the longer window but originate from the lack of temporal context to assess the observed patterns. This was consistent with the behavior of the TA classifier that improved HIE sensitivity uniformly across labor (Fig 3). Although TLO improved HIE sensitivity, the magnitude of the added improvement was smaller than for the window effect alone. Therefore, using a longer window was the dominant factor for improving the early identification of fetuses at risk of HIE, while indexing TLO further improved sensitivity at all landmarks at once.

There are two potential mechanisms behind the added advantage of TLO, although they cannot be distinguished from our data. First, indexing TLO allows the classifiers to differentially weight the same CTG patterns depending on when in labor they occur. The time-resolved importance trajectories in Fig 5 are consistent with this, as different features reached importance peaks at different times. Second, labor duration may be the determining factor for improved HIE sensitivity. TLO is correlated with the length of labor, which is significantly different across groups (Table 2). Thus, the model could learn that after several hours in labor, the risk of HIE increases sharply and independently of CTG patterns. Such behavior cannot be analyzed from the importance trajectories, as TLO was held fixed within bins and we could not quantify its time-resolved importance. While the dominating mechanism behind the advantage of TLO cannot be established from our experiments, prolonged labor is a known risk factor of HIE [30,31]. An important consideration is that the time axis in labor, and TLO specifically, are strongly correlated with all CTG features [22,23]. Thus, the TI classifier could be exploiting that correlation to infer the elapsed time in labor from the ensemble of CTG features when TLO was not available. Indexing TLO made the task much easier for the classifier, and allowed it to focus more on finding discriminative patterns across features rather than inferring TLO. This could explain why the importance of some CTG features appeared to shift towards TLO in the TA classifier, while the overall performance of the classifier improved significantly (Table 6).

Whichever mechanism dominated, TLO is only available when labor onset was identified. In our dataset, labor onset could not be determined in 22,321 births, which represented 10.7% of records with extracted FHR features. Given the size of our cohort, exclusion of these infants was preferred to ensure confidence in the TLO variable. In other smaller cohorts, this cost may be prohibitive. Thus, the choice between TI and TA models is not purely about performance but about data availability. Systems that aim to adopt our approach need a reliable and automated method for determining labor onset, retrospectively for training [24], and prospectively for deployment. Our results showed that accounting for TLO consistently improved the early identification of infants at risk of HIE; whenever labor onset can be reliably identified, TLO should be included in future models.

### 4.3 Clinical lead time and alarm burden

For an intrapartum decision support system to be clinically useful, alarms must be triggered with a longer lead time than the time required to act. In general, emergency Caesarean deliveries can be completed in 30 min from decision to delivery [20,21]. This is why we ignored the last two epochs (40 min) before delivery; alarms triggered after that point may not enable preventive interventions. In our study, the 1-hour TI classifier had the best HIE and acidosis sensitivity at the actionable deadline. However, this classifier was consistently inferior at the 3 h and 6 h landmarks. Regardless of how impressive the performance towards the end of labor may be, the detected cases are clinically unrecoverable. Thus, we argue that, for the development of these systems, detection lead time should be an important optimization target as well as maximum sensitivity.

Another important consideration is the number of alarms triggered and the risk of alarm burden. Alerts at FPR = 15% represent a risk flag, not the decision for operative delivery. These alerts identify which labors warrant closer clinical evaluation. Clinical actions are graduated and do not start with immediate Caesarean delivery: clinicians may increase the frequency of reviews, obtain advice from senior staff, or attempt intrauterine resuscitation. Only if the tracing deteriorates further should there be preparation for expedited delivery [32]. The value of early identification lies in allowing enough time for the graduated responses to be applied in an anticipatory manner rather than a reactive manner. Moreover, our system allows for this by design: it outputs continuous scores through labor, so clinicians can track risk as it evolves and support their decisions on that. Interestingly, Fig 4 shows that the FPR was different for the two healthy groups. At the actionable deadline, the FPR in the healthy without BG group was 12.0%, whereas for the healthy with BG group was 28.7%. While the aggregated FPR was held stable at 15%, these within-class differences show the effect of selecting for BG testing in the data distribution. The healthy group without BG had shorter and less worrisome labors, and it drove the total FPR down as the majority component of the healthy class. In contrast, the healthy group with BG data presented some worrying patterns that prompted clinicians to perform the biochemical assessment; it is likely that these are the same worrying patterns that caused the higher alert rate by our classifier. In clinical practice, these cases would prompt closer clinical evaluation, even when the outcomes were healthy.

The NNE quantified the operational cost of each true detection. The TA model triggered 161.5 alarms for each identified HIE case at the actionable deadline (S2 Table). The NNE was lower earlier in labor, 128.9 at 3 h and 111.1 at 6 h before delivery. Also, when considering the incidence of HIE, it is evident that the achieved NNE for HIE is much lower than that obtained if the full cohort were evaluated (NNE = 573.0). Thus, a high NNE is unavoidable given the low incidence of HIE; the NNE for acidosis was far lower (24.3) because its incidence is 8.7 times higher than HIE. Thus, NNE is bounded by both model quality and class prevalence and cannot be directly compared across outcomes. Sensitivity remains the comparable metric, and it favored HIE for a stable FPR.

Despite the relatively high NNE, in the context of intrapartum evaluations, we consider it conservative. Non-reassuring and indeterminate tracings in CTG are present at any point in over 80% of intrapartum tracings under the current visual interpretation guidelines [6]. In our cohort, and considering that all HIE cases were detected, this number of alerts would still represent a NNE *>* 450. Thus, a system that flags only 15% of labors is significantly more specific than existing practice and it manages to flag pathological cases much earlier than delivery. Thus, the additional HIE cases identified by the 18-hour TA model were net gains and not the by-product of a more lenient alarm system.

The clinical gap of early identification of HIE risk has also been documented externally. In a 20-year cohort that included 314 HIE cases, half of the HIE cases showing normal CTG tracings at admission were defined as potentially avoidable with appropriate intrapartum care; this corresponded to 22% of all term HIE [33]. Those cases had fewer identifiable antenatal risk factors than the remainder, so their timely identification depends strongly on improved intrapartum monitoring besides risk stratification at admission. Importantly, Table 2 shows that in our cohort 43.4% HIE cases underwent Caesarean delivery for fetal intolerance. All these infants still developed HIE, suggesting that interventions were already too late. Our system addresses this clinical gap, where model selection was performed for early sensitivity, and where over 40% of HIE cases were identified 3 h or longer before delivery.

### 4.4 Comparison with previous work

Table 1 compares the sensitivity reported by other models for a FPR = 15%: Petrozziello et al. reported 53%, McCoy et al. 70%, M’Barek et al. 42–62%, and Mendis et al. 50%. Some of these sensitivities are above our HIE sensitivity at the actionable deadline (55.7% ± 0.8%). However, these numbers are not directly comparable to our results for three main reasons. First, the outcome was different; these models targeted cord-gas acidosis using pH thresholds in the 7.05–7.20 range. In contrast, we targeted severe acidosis and chart-confirmed HIE. These are different outcomes with different temporal structures. Second, HIE prevalence is ∼0.17%, far less prevalent than the target class in the comparator studies. Also, our cohort did not select for the most extreme cases. In comparison, one comparator excluded cases with intermediate 7.05 ≤ pH *<* 7.15 from both training and evaluation [10], which completely changes the FPR axis upon which the operating point is selected and removes the cases in which discrimination is hardest. Finally, these sensitivities were reported at a single retrospective point in time, the time of delivery, whereas ours are reported continuously and for several hours before delivery.

Another difference between our results and previous reports is the way the sensitivities were computed in the test set. The studies in Table 1 read the model sensitivity at a target FPR directly from the test-set ROC curve. This implies an operating point that is selected with knowledge of the test set score distributions. In contrast, in our study, we fixed *τ* on inner cross-validation predictions and applied it unchanged to the test set predictions. This is the only way that alarms can be triggered in deployment, where the distribution of scores for new infants is unknown. This approach is also harder to report, as nothing guarantees that the transferred threshold will maintain a stable FPR. In our study, this stability was achieved, and FPR was kept at a nominal 15% across all models and all folds. The only exception to FPR stability occurred when we simulated the deployment of a model trained and calibrated only in the last hour of labor, where FPR rose to 16.2% (Table 5). This underscores the importance of calibrating for the intended time window in which the system will operate. More importantly, comparisons of sensitivities at fixed FPR should state explicitly whether the operating point was selected on test data or transferred from a previous validation procedure. This distinction is crucial for assessing generalizability of a system and prospective stability of its alarm burden.

### 4.5 Limitations and future work

Despite its merits, our study also carries limitations that must be discussed. While it proved beneficial, extending the training window unavoidably increases label uncertainty. Early epochs of a pathological case may be physiologically healthy, which implies that the training set contains mislabeled epochs. While the increase in HIE sensitivity indicates that label noise did not dominate model performance, it still could not be eliminated. There is a second source of label uncertainty in our cohort: 139,512 out of 171,246 controls were classified as healthy despite not having blood-gas data to confirm it. Some of these infants may have had unmeasured acidosis, but presented no further complications at birth. Such cases could potentially bias FPR upwards; alerts on these infants are scored as false positives. Conversely, the confirmed acidosis cases may present overt CTG abnormality, since only clinician suspicion triggered BG testing, which would bias model sensitivity upward.

Regarding the generalizability of our results, all our data came from a single health system and was evaluated retrospectively. Although we included a total of 15 KPNC hospitals, all of them are part of the same organization, using similar systems and protocols. Thus, external validation on other populations and with different monitoring equipment is desirable. Unfortunately, no long-term open database of confirmed HIE cases currently exists, which is an important gap in the field. Moreover, our conclusions are based on retrospective data, which cannot account for clinician actions in response to early alerts. Whether early sensitivity performance would hold in clinical deployment, and whether early alerts would translate into improved outcomes of labor requires prospective evaluation.

Another caveat is that our framework was optimized for chronic, progressive deterioration. HIE caused by sentinel events such as cord prolapse, uterine rupture, or placental abruption may lack discriminating FHR signs before the event and may benefit less from an extended window. Also, the permutation importance analysis should be read as an assessment of association and not causation. Our time-resolved trajectories are descriptive of model behavior in time, and model dependence, but they do not determine causal pathways in the pathophysiology of HIE. Thus, these results should not be used to infer physiological processes.

This study has also revealed further gaps that could be addressed by future studies. Here, we outline some of them:

- **Model Architecture:** our study investigated a property of the training data, which is independent of model architecture. Thus, we held the architecture fixed by design and alarms depended only on single epochs. Model encoders could expand the feature extraction process, such that the model learns the relevant information directly from the data and does not depend on handcrafted features [12]. Similarly, sequence classifiers like BiLSTM or Transformers could be used to model the temporal behavior of the CTG properties and determine when a trajectory is moving away from what is expected for a healthy outcome [34]. Thus, a future study could investigate the architecture best suited to exploit an extended window to improve early HIE sensitivity.
- **Target Expansion and Unlabeled Data:** while we provided measures of sensitivity as a function of time before delivery, our classifier was still only a binary classifier. Future studies could explore predicting time-to-event besides risk of event alone. This could provide tangible information to clinicians to plan management. Similarly, severity grading could also be modeled. Due to the extremely low incidence of HIE, we grouped together all categories of encephalopathy. An expanded cohort, or a multi-center study, could disaggregate cases by severity of neurological injury to also provide further information to clinicians that support their decisions in the delivery room. Finally, there is a need for better utilization of unlabeled cases. In our study, all data that did not include BG data was labeled as healthy provided that they presented no complications at birth. However, other studies have opted for full exclusion of these cases [11], which discards valuable clinical data. Novel methods, such as self-supervised learning, provide alternatives that take advantage of unlabeled signals for representation learning (encoder pretraining), which ensures that all data can be included in the development of novel models [35].

## Conclusion

FHR patterns evolve in labor and with the development of acidosis and subsequent HIE. For AI systems to be able to track these changes, and detect infants at high risk as early as possible, they must account for this evolution. Expanding the analysis window, from the standard final hour of labor, and indexing TLO into the classifiers, significantly improve their early warning capabilities. Thus, it is necessary to treat labor as a nonstationary process and develop time-aware models to enhance the early identification of fetuses at high risk of intrapartum HIE.

## Data Availability

The datasets generated and analyzed during the current study are not publicly available due to patient privacy policies. Unidentified cardiotocography (CTG) records may be available for research but require submitting a study proposal. The CTG data cannot leave KPNC firewalls under any circumstances. Interested parties might follow these steps: (1) submit a proposal to KPNC Division of Research that includes the author MK (or his designate) as a co-investigator, (2) the proposal will be reviewed and approved by the KPNC Institutional Review Board for the Protection of Human Subjects, and (3) a Data Use Agreement between the interested parties and the Division of Research will be executed. The interested parties must commit to research use only, with no commercial use of the datasets.

## Acknowledgments

We are grateful for the administrative support provided by: Brigid Acuna, Jennifer Baker, and Eileen Walsh. We also want to extend our deep appreciation for the advice provided by Dr. Emily Hamilton in the conception and writing of this study.

## Supporting information

**S1 Table. Performance across the varying training window length for the time-invariant classifier.** Sensitivity, NNE, and FPR for the acidosis and HIE groups, for each training window *W* ∈ {1, 3, 6, 9, 12, 18, 24, 36, 48, 72} h. All metrics but FPR are reported at the actionable deadline (40 min), and at 3 h and 6 h before delivery. *W ^∗^* = 18 h was selected by the step-down procedure where *W*_peak_ = 72 h was the best performer. Significance was evaluated against *W*_peak_ by one-sided Wilcoxon signed-rank test with Holm-Bonferroni correction across all evaluated *W*. Supports Section 3.2 in the main text.

**S2 Table. Continuous deployment performance and comparison of the 1-hour TI, 18-hour TI, and 18-hour TA classifiers.** The table reports sensitivity, NNE, AUROC, and FPR for the HIE and acidosis groups at the actionable deadline (40 min), 3 h, and 6 h before delivery, computed on the full test set. The lower block reports the three pairwise differences among the three models. Two-sided Wilcoxon signed-rank test with Holm-Bonferroni correction across all compared metrics. The best value for each metric is bold. Differences were computed per fold, averaged, and then rounded, so they may differ by ∼ 0.1. Supports Section 3.3.

**S3 Table. Global permutation importance for the 1-hour and 18-hour time-invariant classifiers.** The 15 features with the highest importance are shown. The importance was quantified as the percentage drop in infant-level partial AUROC (0–15% false positive rate) when the feature is permuted, for *W* = 1 h and *W ^∗^* = 18 h. Effect size is the rank-biserial correlation, the *p*-values are from two-sided Wilcoxon signed-rank tests with Holm-Bonferroni correction across all the 40 features. Supports Section 3.4.

## References

1. Lee AC, Kozuki N, Blencowe H, Vos T, Bahalim A, Darmstadt GL, et al. Intrapartum-related neonatal encephalopathy incidence and impairment at regional and global levels for 2010 with trends from 1990. Pediatric Research. 2013;74(1):50–72. doi:10.1038/pr.2013.206.

2. Janbu T, Nesheim BI. Uterine artery blood velocities during contractions in pregnancy and labour related to intrauterine pressure. BJOG: An International Journal of Obstetrics & Gynaecology. 1987;94(12):1150–5. doi:10.1111/j.1471-0528.1987.tb02314.x.

3. Lear CA, Wassink G, Westgate JA, Nijhuis JG, Ugwumadu A, Galinsky R, et al. The peripheral chemoreflex: indefatigable guardian of fetal physiological adaptation to labour. The Journal of Physiology. 2018;596(23):5611–23. doi:10.1113/JP274937.

4. Greco P, Nencini G, Piva I, Scioscia M, Volta C, Spadaro S, et al. Pathophysiology of hypoxic–ischemic encephalopathy: a review of the past and a view on the future. Acta Neurologica Belgica. 2020;120(2):277–88. doi:10.1007/s13760-020-01308-3.

5. Farquhar CM, Armstrong S, Masson V, Thompson JM, Sadler L. Clinician identification of birth asphyxia using intrapartum cardiotocography among neonates with and without encephalopathy in New Zealand. JAMA Network Open. 2020;3(2):e1921363. doi:10.1001/jamanetworkopen.2019.21363.

6. Jackson M, Holmgren CM, Esplin MS, Henry E, Varner MW. Frequency of fetal heart rate categories and short-term neonatal outcome. Obstetrics & Gynecology. 2011;118(4):803–8. doi:10.1097/AOG.0b013e31822f1b50.

7. Alfirevic Z, Gyte G, Cuthbert A, Devane D. Continuous cardiotocography (CTG) as a form of electronic fetal monitoring (EFM) for fetal assessment during labour. Cochrane Database of Systematic Reviews. 2017;(2). doi:10.1002/14651858.CD006066.pub3.

8. Cornet MC, Kuzniewicz M, Scheffler A, Forquer H, Hamilton E, Newman TB, et al. Perinatal hypoxic-ischemic encephalopathy: incidence over time within a modern US birth cohort. Pediatric Neurology. 2023;149:145–50. doi:10.1016/j.pediatrneurol.2023.08.037.

9. Lovers A, Daumer M, Frasch MG, Ugwumadu A, Warrick P, Vullings R, et al. Advancements in fetal heart rate monitoring: a report on opportunities and strategic initiatives for better intrapartum care. BJOG: An International Journal of Obstetrics & Gynaecology. 2025;132(7):853–66. doi:10.1111/1471-0528.18097.

10. Petrozziello A, Redman CW, Papageorghiou AT, Jordanov I, Georgieva A. Multimodal convolutional neural networks to detect fetal compromise during labor and delivery. IEEE Access. 2019;7:112026–36. doi:10.1109/ACCESS.2019.2933368.

11. McCoy JA, Levine LD, Wan G, Chivers C, Teel J, La Cava WG. Intrapartum electronic fetal heart rate monitoring to predict acidemia at birth with the use of deep learning. American Journal of Obstetrics and Gynecology. 2025;232(1):116.e1-116.e9. doi:10.1016/j.ajog.2024.04.022.

12. M’Barek IB, Jauvion G, Merrer J, Koskas M, Sibony O, Ceccaldi PF, et al. DeepCTG® 2.0: Development and validation of a deep learning model to detect neonatal acidemia from cardiotocography during labor. Computers in Biology and Medicine. 2025;184:109448. doi:10.1016/j.compbiomed.2024.109448.

13. Mendis L, Karmakar D, Palaniswami M, Brownfoot F, Keenan E. Fusing Tabular Features and Deep Learning for Fetal Heart Rate Analysis: A Clinically Interpretable Model for Fetal Compromise Detection. IEEE Transactions on Biomedical Engineering. 2026. doi:10.1109/TBME.2026.3652309.

14. Comert Z, Şengür A, Budak U, Kocamaz AF. Prediction of intrapartum fetal hypoxia considering feature selection algorithms and machine learning models. Health Information Science and Systems. 2019;7(1):17. doi:10.1007/s13755-019-0079-z.

15. Low JA. Intrapartum fetal asphyxia: definition, diagnosis, and classification. American Journal of Obstetrics and Gynecology. 1997;176(5):957–9. doi:10.1016/S0002-9378(97)70385-5.

16. Yeh P, Emary K, Impey L. The relationship between umbilical cord arterial pH and serious adverse neonatal outcome: analysis of 51 519 consecutive validated samples. BJOG: An International Journal of Obstetrics & Gynaecology. 2012;119(7):824–31. doi:10.1111/j.1471-0528.2012.03335.x.

17. Malin GL, Morris RK, Khan KS. Strength of association between umbilical cord pH and perinatal and long term outcomes: systematic review and meta-analysis. BMJ. 2010;340:c1471. doi:10.1136/bmj.c1471.

18. Rieger B, Iqbal S, Garavatti E, Shen J, Morphew T, Inder TE. Evaluating the Predictive Value of Umbilical Cord and Infant Blood Gases in Hypoxic-Ischemic Encephalopathy. The Journal of Pediatrics. 2026. doi:10.1016/j.jpeds.2026.115148.

19. Dellinger EH, Boehm FH, Crane MM. Electronic fetal heart rate monitoring: early neonatal outcomes associated with normal rate, fetal stress, and fetal distress. American Journal of Obstetrics and Gynecology. 2000;182(1):214–20. doi:10.1016/S0002-9378(00)70515-1.

20. Helmy W, Jolaoso A, Ifaturoti O, Afify S, Jones M. The decision-to-delivery interval for emergency caesarean section: is 30 minutes a realistic target? BJOG: An International Journal of Obstetrics & Gynaecology. 2002;109(5):505–8. doi:10.1016/S1470-0328(02)00491-3.

21. Thomas J, Paranjothy S. The national sentinel caesarean section audit report; 2001. Available from: https://orca.cardiff.ac.uk/id/eprint/93112.

22. Vargas-Calixto J, Wu YW, Kuzniewicz M, Cornet MC, Forquer H, Gerstley L, et al. Time-dependent Association Between Cardiotocographic Features and Hypoxic-ischemic Encephalopathy. IEEE Access. 2025. doi:10.1109/ACCESS.2025.3570371.

23. Vargas-Calixto J, Kuzniewicz MW, Cornet MC, Wu YW, Lahiri A, Gerstley L, et al. Time-surrogate variables enhance the association between cardiotocographic features and intrapartum hypoxic-ischemic encephalopathy. PLOS Digital Health. 2026 08;5(8):1-24. doi:10.1371/journal.pdig.0001111.

24. Kearney RE, Wu YW, Vargas-Calixto J, Kuzniewicz MW, Cornet MC, Forquer H, et al. Construction of a comprehensive fetal monitoring database for the study of perinatal hypoxic ischemic encephalopathy. MethodsX. 2024;12:102664. doi:10.1016/j.mex.2024.102664.

25. Cornet MC, Kuzniewicz MW, Scheffler AW, Garabedian C, Gaw SL, Wu YW. Maternal fever during labor and the risk of neonatal encephalopathy: duration and magnitude of hyperthermia. American Journal of Obstetrics and Gynecology. 2025. doi:10.1016/j.ajog.2025.07.046.

26. Warrick P, Hamilton E, Macieszczak M. Neural network based detection of fetal heart rate patterns. In: Proceedings. 2005 IEEE International Joint Conference on Neural Networks, 2005.. vol. 4. IEEE; 2005. p. 2400–5. doi:10.1109/IJCNN.2005.1556278.

27. Warrick PA, Hamilton EF. Antenatal fetal heart rate acceleration detection. In: 2016 Computing in Cardiology Conference (CinC). IEEE; 2016. p. 893–6.

28. Janitza S, Celik E, Boulesteix AL. A computationally fast variable importance test for random forests for high-dimensional data. Advances in Data Analysis and Classification. 2018;12(4):885–915. doi:10.1007/s11634-016-0276-4.

29. Lear CA, Beacom MJ, Dhillon SK, Lear BA, Mills OJ, Gunning MI, et al. Dissecting the contributions of the peripheral chemoreflex and myocardial hypoxia to fetal heart rate decelerations in near-term fetal sheep. The Journal of Physiology. 2023;601(10):2017–41. doi:10.1113/JP284286.

30. Reynolds AJ, Murray ML, Geary MP, Ater SB, Hayes BC. Uterine activity in labour and the risk of neonatal encephalopathy: a case control study. European Journal of Obstetrics & Gynecology and Reproductive Biology. 2022;274:73–9. doi:10.1016/j.ejogrb.2022.05.011.

31. Torbenson VE, Tolcher MC, Nesbitt KM, Colby CE, El-Nashar SA, Gostout BS, et al. Intrapartum factors associated with neonatal hypoxic ischemic encephalopathy: a case-controlled study. BMC Pregnancy and Childbirth. 2017;17(1):415. doi:10.1186/s12884-017-1610-3.

32. Mercer L, Caughey AB, Raghuraman N, Saposnik SR. ACOG Clinical Practice Guideline No. 10: Intrapartum fetal heart rate monitoring: Interpretation and management. Obstetrics & Gynecology. 2025;146(4):583–99.

33. Tarvonen M, Jernman R, Stefanovic V, Tuppurainen V, Karikoski R, Haataja L, et al. Hypoxic-ischemic encephalopathy following intrapartum asphyxia: is it avoidable? American Journal of Obstetrics and Gynecology. 2025. doi:10.1016/j.ajog.2025.04.073.

34. Khan MJ, Vatish M, Davis Jones G. PatchCTG: A patch cardiotocography transformer for antepartum fetal health monitoring. Sensors. 2025;25(9):2650. doi:10.3390/s25092650.

35. Vargas-Calixto J, Nateghi M, Ward J, Robertson J, Sebastĩao Y, Patterson J, et al. Self-Supervised Representation Learning of Fetal Heart Rate Outperforms a Foundation Model in Identifying Fetal Acidosis. In: 2026 Computing in Cardiology (CinC). Madrid, Spain; 2026.

